# Magnetic Resonance Imaging Data Phenotypes for the Parkinson’s Progression Markers Initiative

**DOI:** 10.1101/2024.09.23.24313179

**Authors:** Brian B Avants, Leon Fonville, Olivia Hampton, Alexandra Reardon, Andrew Stenger, Xue Wang, Nicholas J Tustison, James R Stone, Philip A Cook, Barbara Marebwa, Lana M Chahine, Kathleen L Poston, Kenneth Marek, Lino Becerra, Roger Gunn, the Alzheimer’s Disease Neuroimaging Initiative

## Abstract

The Parkinson’s Progression Markers Initiative (PPMI) delivers multiple modality MRI (M3RI) and biomarker data for a comprehensive longitudinal study of Parkinson’s Disease (PD). These provide quantitative indices of deep brain and cortical structure (T1-weighted MRI), microstructural integrity of brain tissue (diffusion-weighted imaging) and resting brain function (resting state functional MRI). Integrating and uniformly analyzing M3RI alongside non-imaging biological and clinical data is challenging due to the distinct nature of each modality. This study systematically organizes these complex data into a structured format, provides a PD-focused evaluation of the methodologies and evidence for technical robustness of the approach. The cohort encompasses 841 idiopathic PD, 309 genetic PD, 1364 presymptomatic PD and 240 control subjects at baseline with followup at a mean of 1.83 years.

## Background & Summary

Parkinson’s Disease (PD) is characterized by the progressive accumulation of Lewy bodies, primarily composed of misfolded alpha-synuclein, and appearing in the substantia nigra at an early stage (Fearnley and Lees 1991). The spread of this pathology correlates with both motor and non-motor symptoms of PD, underscoring alpha-synuclein’s pivotal role in disease progression (Lee and Trojanowski 2006; Dickson et al. 2009; Calabresi et al. 2023). The synuclein amplification assay (SAA) significantly advanced PD research by enabling in vivo confirmation for the first time. The Parkinson’s Progression Markers Initiative (PPMI) study enhances this development by offering a comprehensive dataset of subjects assessed with SAA and multimodal MRI (M3RI), facilitating the monitoring of PD progression and the impact of synucleinopathy on brain structure and function (Shahnawaz et al. 2020; Siderowf et al. 2023).

Analyzing longitudinal M3RI in the PPMI study necessitates accessible data representations for the PD research community. While each MRI modality provides distinct insights, their combined analysis is challenging due to the data’s high dimensionality and computational demands. Therefore, creating clear, accessible data representations is crucial for advancing PD research and fostering new discoveries in particular using multi-view data linking evidence of pathology, symptoms and imaging (Nemmi et al. 2019; Tremblay et al. 2020; Markello et al. 2021).

The study utilizes the Advanced Normalization Tools X (ANTsX) ecosystem to process PPMI MRI from 2010 to early 2024, focusing on T1-weighted, diffusion-weighted, and resting-state functional rsfMRI. The extended duration of data collection underscores the necessity for robust processing techniques that manage the resultant heterogeneity in images. ANTsX, leveraging decades of MRI analysis expertise, employs advanced techniques and integrates open science, deep learning, and machine learning for efficient multi-site M3RI data processing (B. B. Avants et al. 2015; Stone et al. 2020; Tustison et al. 2021)

To establish face validity, this study leverages not only PPMI M3RI, but also the Alzheimer’s Disease Neuroimaging Initiative (ADNI) (Veitch et al. 2024) and traveling subject data in three cohorts (Hawco et al. 2022; Tanaka et al. 2021; Tong et al. 2019). We analyze these data collectively and uniformly to establish reliability benchmarks and to demonstrate feasibility of consistent processing in multi-site studies such as PPMI. Additionally, we exemplify statistical models using PPMI data that are appropriate for unlocking key questions relevant to biomarker-confirmed PD and related conditions.

Historically, MRI research on PD has primarily utilized T1-weighted (T1w) structural imaging to investigate neuroanatomical changes. A consistent finding across these studies has been the identification of early neurodegeneration in mid-brain regions, particularly the substantia nigra (Schwarz et al. 2011; Aquino et al. 2014; Ryman and Poston 2020; Poston et al. 2020). Diffusion-weighted MRI (dwMRI) has further enhanced our understanding by allowing for the investigation of white matter microstructural integrity which may be impacted not only in sporadic PD (Péran et al. 2010; Owens-Walton et al. 2024) but also in genetic PD (Tolosa et al. 2020; Owens-Walton et al. 2024). Similarly, resting state functional MRI (rsfMRI) has unveiled alterations in network connectivity in PD patients, highlighting changes in the functional integration and segregation of brain networks involved in motor and cognitive functions (Hacker et al. 2012; Kim et al. 2017; Esposito et al. 2013). Taken together, these findings suggest that PD impacts both the structural and functional aspects of the brain. More integrative research in PD (Menke et al. 2009; Markello et al. 2021) is needed to determine the sequence of these changes and how they may relate to alpha-synuclein and potential copathology or comorbidity (Simuni et al. 2024).

The present study extends previous foundations by providing standardized imaging data phenotypes (IDPs) for PPMI with a particular emphasis on an accessible tabular representation. The summary IDPs are computed with ANTsPyMM v1.4.0 and depend on standard anatomical and functional hierarchies that are well-established in the field and consistently integrated in this work across modalities. This approach supports investigations based on T1w structural imaging, dwMRI, and rsfMRI either independently or collectively. Importantly, these imaging variables easily merge with the associated demographics, SAA status, clinical data such as the Movement Disorder Society-Sponsored Unified Parkinson’s Disease Rating Scale (MDS-UPDRS) (Disease 2003) and standard PPMI DAT-SPECT summary measurements (Bega et al. 2021; Droby et al. 2022). Through this integrative methodology, we aim to contribute to a deeper understanding of PD, facilitating the development of more effective diagnostic and therapeutic strategies and accelerating PD research.

## Methods

### MRI data collection

MRI data collection occurred between 2010 and an April 2024 cutoff date for these data. Two phases of MRI collection occurred in PPMI; the first collected T1w and later DTI as part of exploratory investigations. In 2020, a new phase of collection sought to improve both MRI quality and consistency and expand the number of modalities collected. The sequences used at each site are provided in detail at (B. Avants 2024) and in the original Laboratory of Neuroimaging (LONI) source data (described below in Data Records). The “phase” of data collection is captured in the variable `imaging_protocol`. The PPMI data includes control subjects, idiopathic (sporadic) PD subjects and genetic PD subjects characterized by GBA, LRRK2, SNCA and PRKN mutations; the latter two groups appear infrequently in this PPMI M3RI cohort. Additionally, presymptomatic subjects comprise a substantial and growing portion of the cohort; these are also characterized by genetic mutations and/or early risk factors for PD (hyposmia or RBD) (Siderowf et al. 2023). Table 1 summarizes the cohort characteristics. In the following sections, we focus on the PD cohort with SAA measurements as the presymptomatic cohort is undergoing significant additional data collection.

**Table 1.**
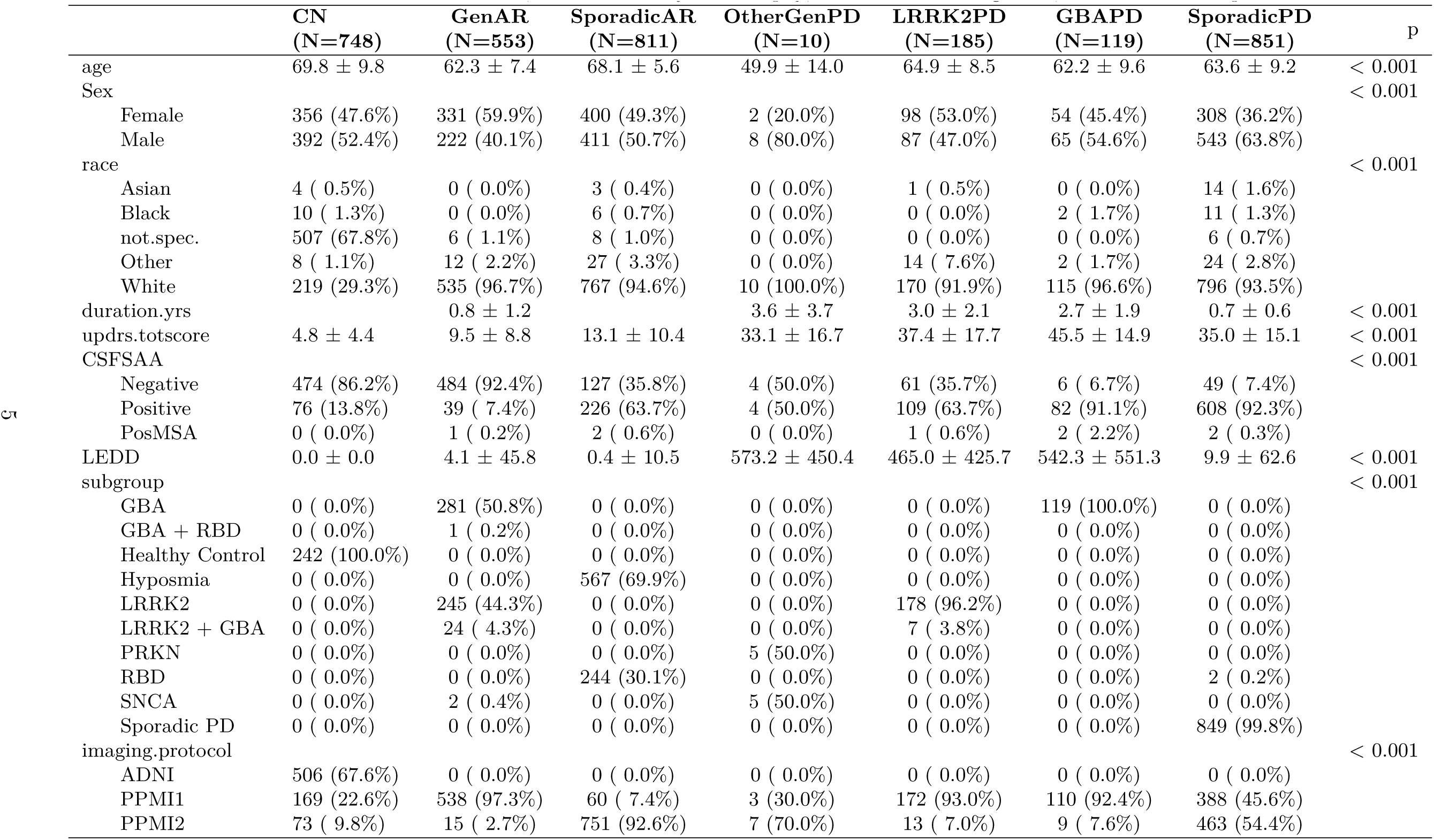
Baseline PPMI IDP cohort. AR=at-risk; MSA = multi-system atrophy; OtherGen = other genetic; RBD = REM Sleep Behavior Disorder.

### Data Organization

Raw DICOM data was downloaded from LONI and converted to nifti format via `dcm2niix` (Li et al. 2016). These data were then organized into a directory tree following the NRG format illustrated in Figure 1. This BIDS-like structure (Gorgolewski et al. 2016) is defined to aid in longitudinal analyses of multiple modality data and intends to support: (a) sortable and specific dates associated with imaging sessions; (b) links between the data on disk and its origin (LONI) through the “Image ID”; (c) easy maintenance of multiple modality data collections; and (d) predictable input/output structure. Critically, the unique ID allows the original data associated with an IDP to be easily found in LONI. In brief, this system assigns each image – and categories of derivative data – a directory and individual file name that assist in making data findable, accessible, interpretable and reproducible (FAIR) for both early and downstream processing.

**Figure 1:**
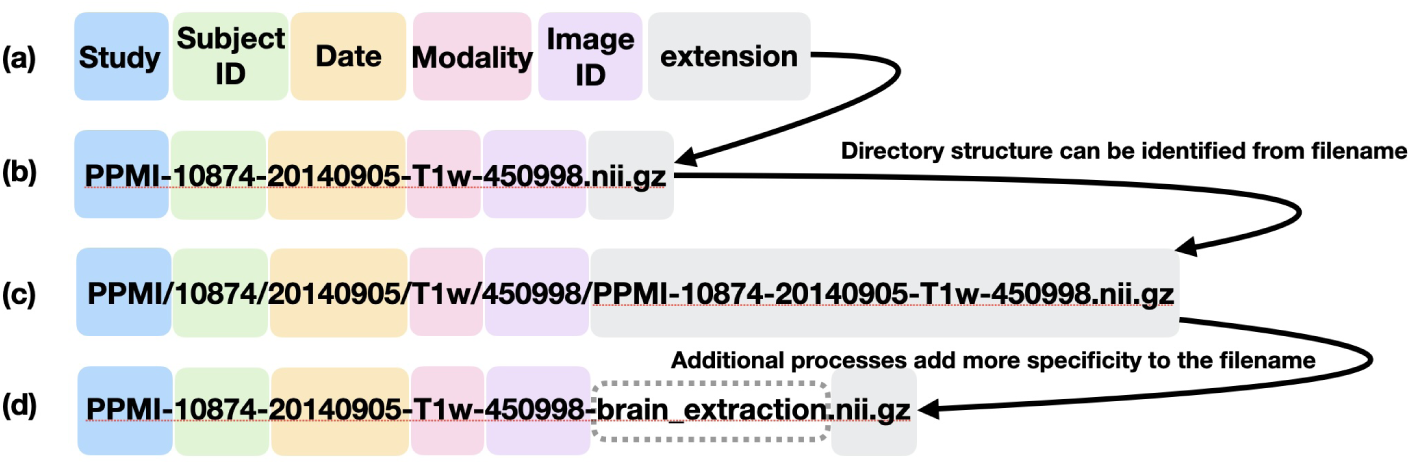
The NRG format supports predictable and interpretable data storage and processing that can easily be tied back to the LONI source dicom. The filename proceeds from less specific information (Project ID at reader’s left) to the most specific (unique image ID at reader’s right). A specific character – here the dash – is reserved exclusively as a separator between the stages of information.

### ANTsPyMM processing

ANTsPyMM collects and documents best ANTsX practices for both data inspection and IDP generation for the modalities of interest in a single python package. While ANTsPyMM supports BIDS format, it behaves most predictably and safely with NRG format. Each “run” of the integrated multiple modality processing encoded by ANTsPyMM is driven by a data frame that defines a multiple modality “collection” of images for a given subject at a given date. There are two key functions that aid users in defining the appropriate input data structure and sending that data to processing. The first is `antspymm.generate_mm_dataframe` which generates the appropriate multiple modality subject dataframe that documents on disk locations for image sets. The resulting dataframe defines the expected input as well as output structures. The second key function runs the multiple modality processing (`antspymm.mm_csv`) based on the multiple modality subject dataframe. The “Usage Notes” section provides more details on this system with an accompanying reproducible example based on freely accessible multiple modality neuroimaging.

### Semi-automated quality assessment

ANTsPyMM’s primary goal is reliable M3RI IDP generation. Of necessity, it also addresses quality control (QC) with particular focus on the T1w modality i.e. the core anatomical image that represents the most consistently collected MRI in PPMI. T1w is the focus of QC because ancillary modality processing depends heavily on anatomical labels (e.g. tissue segmentation, cortical parcellation) derived from these images. As such, we developed an automated (deep learning based) T1w reviewer that is trained on human (BA) QC reviews. Each T1w image is therefore reviewed internally in the first stage of ANTsPyMM processing by this `resnetGrader` (a deep learning model trained to predict image quality) (B. Avants et al. 2023). The grader will abort processing if the T1w does not achieve a given baseline level of quality. Human visual inspection was performed on images that pass the grader by BA and serves as a sanity check to the automated method. The `resnetGrader` successfully filtered unusable data and we selected a quality cutoff at 1.02 to filter out low quality images. Similarly, the rsfMRI and DTI were reviewed in post hoc fashion. This process involved visually inspecting each estimated FA image and each estimated default mode network connectivity map and its associated mean BOLD image. Particular focus was paid to cases with high motion and/or low SNR; such images were excluded from statistical analyses.

### Neuroanatomical coordinate systems

The statistical interpretation of processed images is aided by automatic anatomic labeling with pre-specified coodinate systems or maps overlaid on each subject’s neuroimage. We leverage a recent homotopic parcellation (Yan et al. 2023), the Desikan-Killiany-Tourville (DKT) system (Klein and Tourville 2012), the CIT168 atlas (Pauli, Nili, and Tyszka 2018), the Johns Hopkins University (JHU) white matter labels (Mori, Oishi, and Faria 2009), the Schmahmann cerebellar parcellation (Lyu et al. 2024; Carass et al. 2018), brain stem labels (Iglesias et al. 2015), a medial temporal lobe schema (Rizvi et al. 2023) and labels derived from probabilistic maps of the basal forebrain (Liu et al. 2015; Zaborszky et al. 2008). These systems are described in detail in online data dictionary and associated documentation for this project. These coordinate system enable PD researchers to interrogate a variety of hypotheses related to, for example, known functional networks, association hubs, cholinergic networks, the striatum or dopaminergic systems.

### T1-weighted MRI processing

T1-weighted MRI processing is described in detail in (Tustison et al. 2023, 2021). This open-source software ecosystem includes tools for image registration, segmentation, and super-resolution (SR) as customized for the human brain. The derived measurements are tabulated by the neuroanatomical coordinates defined above and include cortical and subcortical measurements and morphological measurements of the hippocampus, basal forebrain and cerebellum. The results of this stage are key to consistent processing of rsfMRI and DWI. We provide both original resolution (OR) and SR results as part of this effort. For SR processing of T1w, the network is applied – first – over the whole head T1w image to double resolution along all axes within the brain parenchyma. Otherwise, SR and OR processing are identical. SR training with 3D perceptual losses is documented in the `python` package siq and is based on `tensorflow` implementations of a volumetric deep back projection network (DBPN) (B. Avants et al. 2023; Tustison et al. 2021; Haris, Shakhnarovich, and Ukita 2020). See Figure 2 for examples of these outputs. IDPs derived from the T1 processing are denoted by prefixes `T1w` and `T1Hier`.

**Figure 2:**
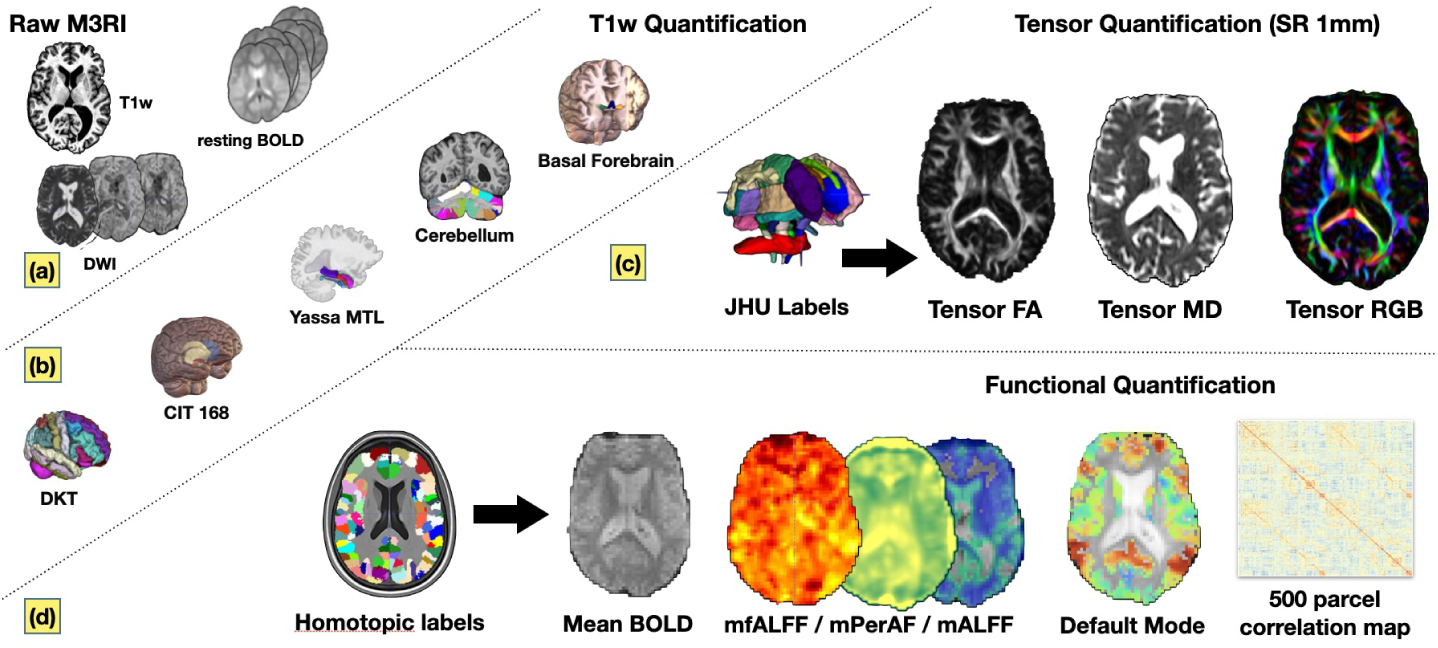
Overview of ANTsPyMM outputs for T1-weighted MRI, diffusion MRI and resting state fMRI. Panel (a) shows example input data; the package does not require all modalities to be present – only T1w. It also handles arterial spin labeling (perfusion), FLAIR and neuromelanin, not covered here. Panel (b) illustrates core T1w outputs across several inter-related and PD relevant systems in the brain. Panel (c) shows the standard outputs associated with DTI. Whole brain tractography is also output but no evaluation results are available to contextualize its performance and, as such, we do not recommend its use. Panel (d) summarizes the various rsfMRI outputs for processing parameter set number 129 referred to with a prefix `rsfMRI_fcnxpro129`.

### Diffusion weighted MRI processing

Diffusion tensor image (DTI) processing leverages best practices from both ANTsX (B. B. Avants et al. 2015; Stone et al. 2023, 2020) and the open-source DTI-focused project DiPy (Garyfallidis et al. 2014). This pipeline is specifically designed to utilize DWI acquisitions with either a single or opposed phase encoding directions. The functionality has been developed to address a broad spectrum of preprocessing requirements, such as motion correction, denoising, dewarping and gradient reorientation, and enhancement through SR techniques, culminating in an optimized DTI reconstruction. The SR stream applies to each volume in the DWI timeseries after motion correction and distortion correction but before tensor fitting i.e. in a relatively minimally invasive fashion. After reconstruction, the pipeline integrates atlas-based labeling and template-based normalization processes, thereby enhancing the anatomical interpretability of the DTI metrics. Figure 4 summarizes the pipeline which follows these steps:

1. **Input Preparation**: The pipeline accepts either a single DWI or a pair of DWI with reversed phase encoding. It also requires associated b-values and b-vectors for each direction, alongside a T1-weighted image and a brain mask for improved spatial accuracy in inter-modality registration.
2. **Initial Reconstruction and Motion Correction**: By default, the DWI data is denoised before performing motion correction. This is skipped when applying SR which integrates denoising. Motion correction aligns DWI volumes within and across acquisitions to a reference mean B0 and mean DWI, reducing artifacts due to subject movement.
3. **Dewarping and Super-Resolution**: Dewarping is applied to correct for distortions between the DWI space and the T1-weighted image. Optionally, SR is applied after dewarping but before the DiPy based reconstruction
4. **Reconstruction of DTI Metrics**: The function employs weighted least squares to reconstruct DTI metrics such as Fractional Anisotropy (FA) and Mean Diffusivity (MD) from the preprocessed DWI data. This step is pivotal in quantifying the diffusion properties of brain tissue.
5. **Atlas-Based Labeling and Registration**: Utilizing the Johns Hopkins University (JHU) atlas and corresponding labels (Mori, Oishi, and Faria 2009), the pipeline performs spatial registration of the DTI to the atlas space. This process facilitates anatomical localization and quantification of DTI metrics within predefined brain regions.
6. **Output Generation**: The pipeline yields a comprehensive output including the reconstructed DTI metrics, summary statistics of these metrics within atlas-defined regions, the spatial registration information, and additional diagnostic metrics such as framewise displacement and signal-to-noise ratio (SNR) assessments spatially and temporally for both B0 and DWI. An example output volumetric tensor image with labels is in Figure 3 and 5.

**Figure 3:**
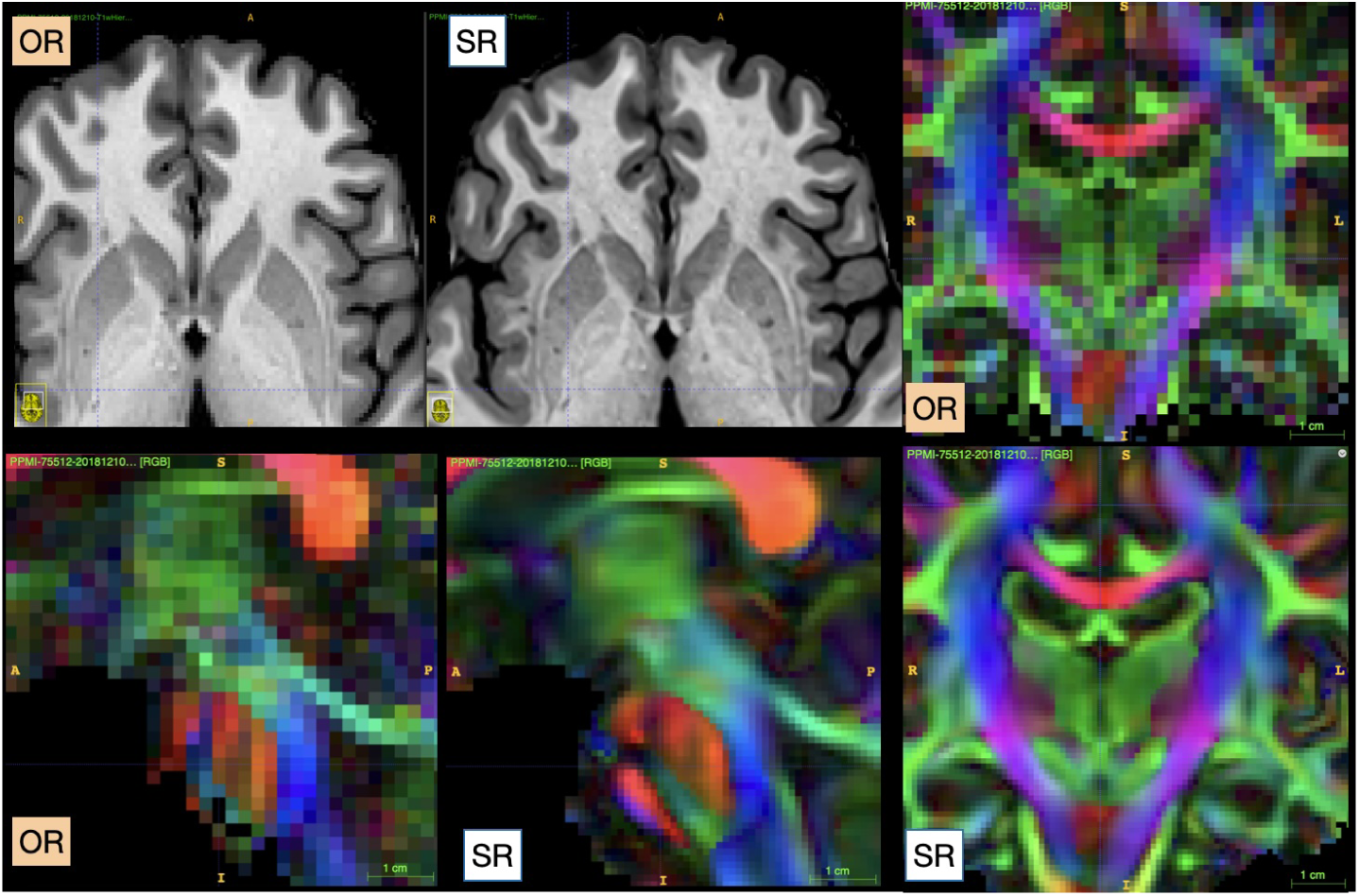
Example ANTsPyMM SR outputs applied to T1-weighted MRI (upper left) and diffusion MRI. T1w is super resolved to 0.5mm isotropic and DTI to 1mm.

**Figure 4:**
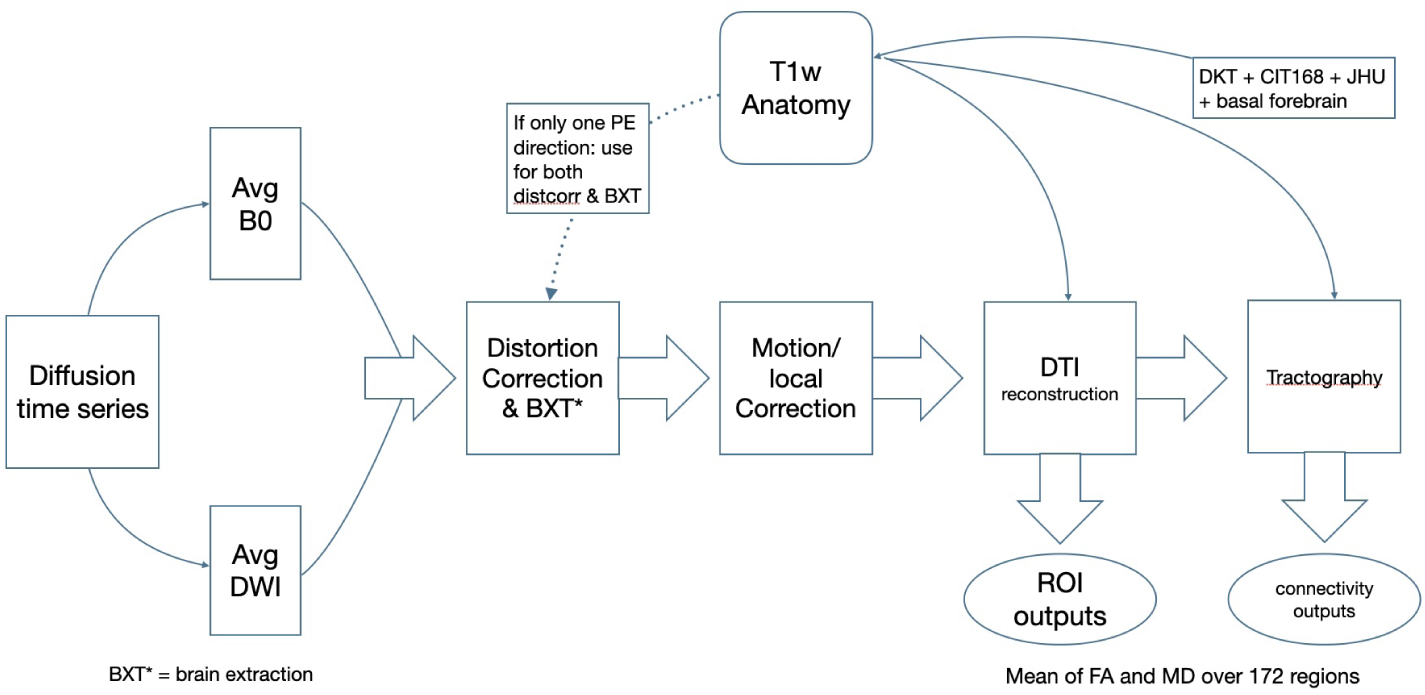
Overview of the DTI processing pipeline based on ANTsX and DiPy. process.

**Figure 5:**
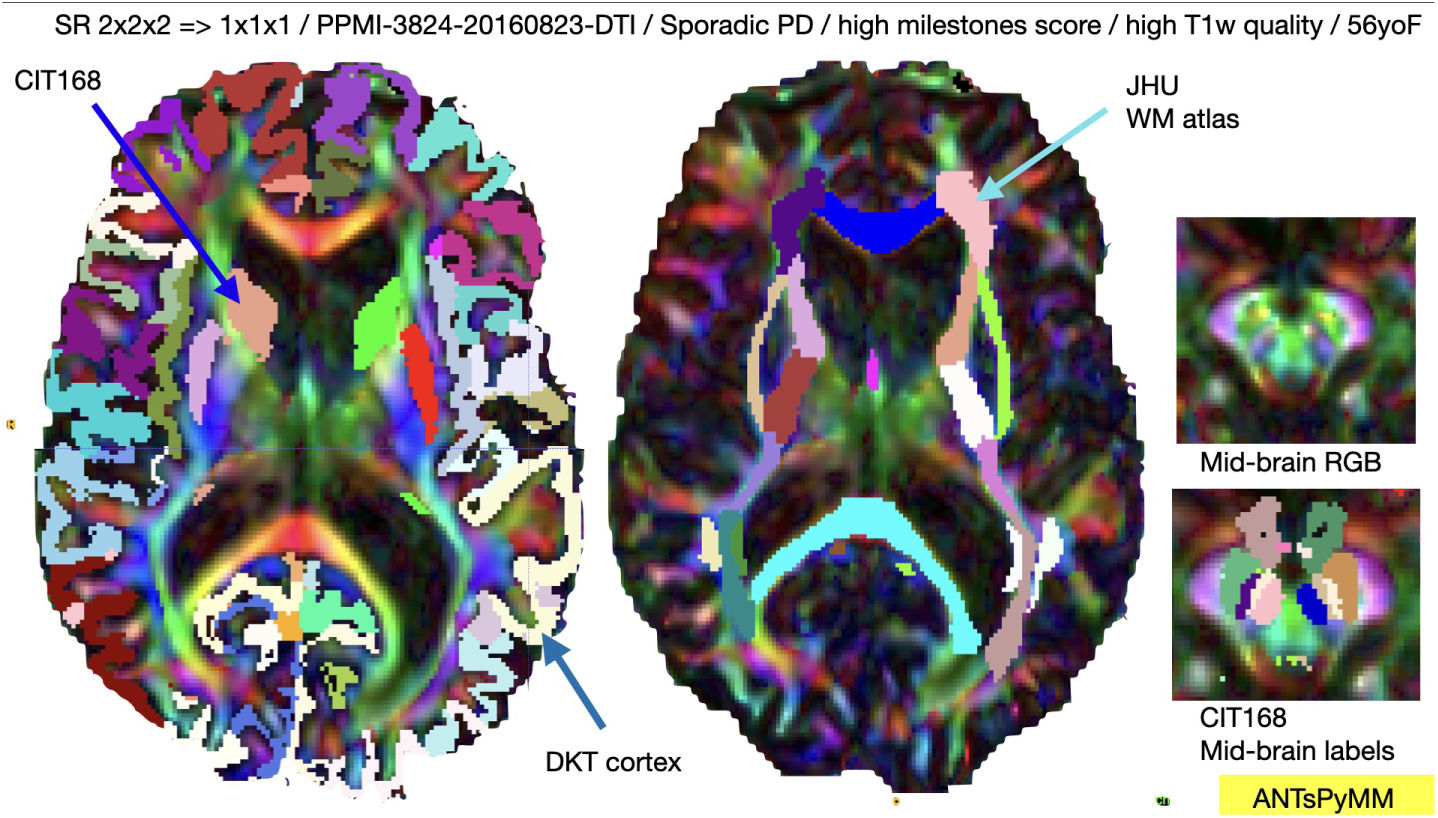
Example SR processing for DTI highligting the multiple coordinate systems to aid interpretation.

IDPs derived from the DWI processing are denoted by prefixes DTI_.

### Resting state functional MRI processing

Resting state functional MRI (rsfMRI) processing builds on prior multi-view M3RI analyses performed in this same ecosystem (B. B. Avants et al. 2015, 2019; B. B. Avants, Tustison, and Stone 2021). The procedure is based on the findings described in three comprehensive evaluation studies (Shirer et al. 2015; Parkes et al. 2018; Noble, Scheinost, and Constable 2019) and is designed to compute both functional activity and correlation maps utilizing the recently proposed homotopic labels to delineate major network systems (Yan et al. 2023). The methodology described below is grounded in contemporary understanding of resting-state fMRI analysis and incorporates recommendations from seminal works regarding optimal preprocessing for minimizing motion artifacts and other sources of noise (Shirer et al. 2015; Parkes et al. 2018). As such, our processing reflects a comprehensive approach to resting-state fMRI IDP extraction for real-world multi-site studies of neurodegenerative disease. Overall, the methods aim to facilitate the reliable extraction of functional connectivity patterns that are consistent with underlying neural mechanisms in PD (Tahmasian et al. 2015; Esposito et al. 2013). Similar to the DWI processing, the procedure accepts either a single image or a pair of images with reversed phase encoding direction. The steps are outlined in Figure 6:

1. **Input Preparation**: Inputs include the raw BOLD fMRI time-series data, a reference volumetric subject-specific fMRI template (automatically generated), and T1-weighted anatomical images all from the same subject. These inputs are foundational for aligning functional data with anatomical landmarks and for ensuring that subsequent analyses are anatomically informed. By default, the input fMRI is upsampled to 3mm isotropic resolution and 8 initial volumes are discarded to allow for both signal and subject stabilization.
2. **Preprocessing**: Initial steps include motion correction, application of a despiking algorithm (a python implementation of AFNI’s 3dDespike (Cox 2012)), and anatomical registration to align the fMRI data with the T1-weighted image. If a pair of images is passed, these same preprocessing steps are applied and results are concatenated along the time axis.
3. **Noise Reduction**: Anatomical CompCor (aCompCor) is used to mitigate physiological and other noise sources. This is based on recommendations from studies examining the impact of preprocessing strategies on functional connectivity (Shirer et al. 2015; Parkes et al. 2018).
4. **Band-pass Filtering and activity calculation**: The application of a specific frequency range for filtering aligns with recommendations from both Shirer et al. (2015) and Parkes et al. (2018), emphasizing the importance of selecting appropriate frequency bands for resting-state analysis. The default frequency bands are based on empirical evaluation studies described below.
5. 5. **Censoring**: Select volumes are censored based on both motion-based and intensity-based outlier detection. The parameters for this stage derive from empirical evaluation studies on public data as discussed below. Both *censored* and *imputed* versions of the time series are created. A summary of censoring results is recorded in several ways but perhaps most relevant are the variables *`minutes_original_data` and *`minutes_censored_data` which provides the length in minutes of the original versus processed data.
6. **Network Correlation Analysis**: This step involves calculating correlation matrices for identified resting-state networks, utilizing labels described above. Both inter and intra-network correlation values are computed for each of the sub-networks provided by the homotopic parcellation.
7. **Functional activity**: is computed with three models: mfALFF, mALFF and mPerAf as described in (Jia et al. 2020). These are versions of fALFF, ALFF and PerAf where each is divided by the global mean in the brain. Summary values are averaged within each of 500 labels in the homotopic label set which facilitates left/right asymmetry and mean values which are critical to studying diseases with laterality effects.

**Figure 6:**
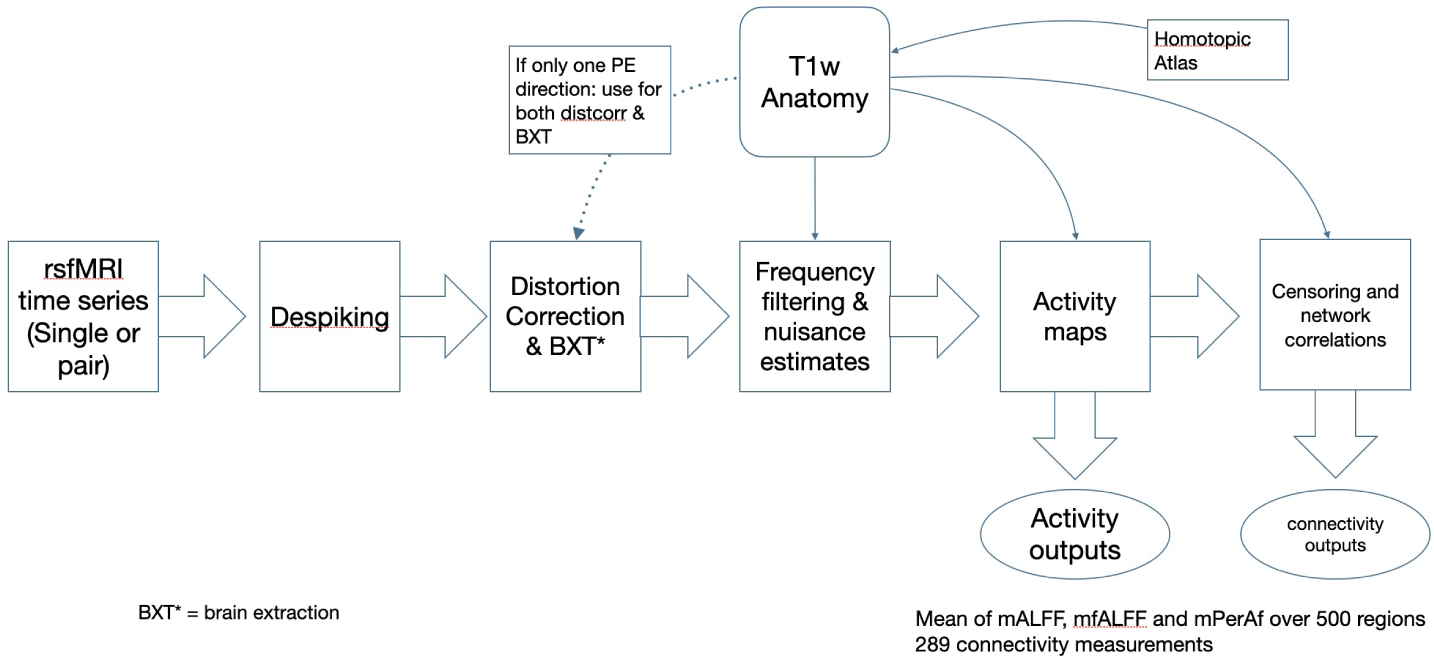
Overview of the rsfMRI processing pipeline based on ANTsX.

Due to the relatively diverse needs of researchers and the variety of rsfMRI that is generally present in public data, we run the above processing with three different sets of parameters. These sets are named by their position in the parameter search data frame as 122, 134 and 129. They encode 3 different choices for outlier rejection (based on motion) and control of nuisance signal via aCompCor. These three parameter choices led to rsfMRI IDPs that were the top performers in terms of reliability and predictive power out of 78 that we tested empirically. See this repository and the technical validation section for further details. IDPs from the rsfMRI processing are denoted by prefixes `rsfMRI_fcnxpro122` for 122 and similarly for 129 and 134.

### Dimensionality reduction with SiMLR

Statistical power in cohorts with diverse composition may be challenged by the number of individual predictors (here, 1178). To address this, we adopt similarity-driven multi-view linear reconstruction (SiMLR) for dimensionality reduction and apply the default settings recommended in prior work (B. B. Avants, Tustison, and Stone 2021; Stone et al. 2023). SiMLR provides a reduced number of predictors by creating “sparse feature sets” that are linked across modalities, allowing for their combined use in joint prediction models. As an unsupervised method, SiMLR identifies a joint low-dimensional space that captures the common variability across these diverse modalities. This enables integrated analyses of MRI and acquired non-imaging data (represented as standard tabular outcomes) within the analytical framework of classical regression. Importantly, this approach can be applied even when only a subset of the variables is present.

Here, the SiMLR decomposition is trained in ADNI subjects and subsequently applied to PPMI thereby decoupling the learning and inference stages. Six matrices were decomposed into 100 joint components. The matrices included one for tabular clinical data and five for neuroimaging including T1w related measurements averaged across left and right, T1w related asymmetry measurements, DTI related measurements averaged across left and right, DTI related asymmetry measurements and resting state connectivity measurements. Although cognition and functionally related clinical scores were employed during decomposition, these were not retained for further application to PPMI. The technical validation section will demonstrate how these learned patterns may be used in analyses integrating PPMI IDPs and clinically relevant metrics both cross-sectionally and longitudinally.

### Data Records

The PPMI IDPs for T1w, rsfMRI and DTI are located at (B. Avants 2024). The neuroimaging and associated standard PPMI demographics and clinical data is hosted in the LONI Imaging Data Archive (LONI IDA). The former is stored in DICOM format and the latter in tabular csv format. Additionally, data dictionaries describing all non-imaging column headers are available on the LONI IDA.

We attach the neuroimaging IDPs to the PPMI Curated Data Cut (v.2024-01-29 `PPMI_Curated_Data_Cut_Public_20240129`) from the LONI IDA. Code for this merging process is within the subtyper package specifically the function merge_ppmi_imaging_clinical_demographic_data. The M3RI IDPs are described in detail here and are in a data table within the `ANTsPyMM` repository (csv format). The full tabular IDPs for both OR and SR outputs are at this location.

We supplement these PPMI data with subjects from ADNI due to the current dearth of control longitudinal neuroimaging in PPMI. As with PPMI, we attach ANTsPyMM IDPs to `ADNIMERGE_10Feb2024` using the `subtyper` function `merge_ADNI_antspymm_by_closest_date`. Select subjects from ADNI are considered for merging with PPMI if they are not diagnosed wtih Alzheimer’s disease or mild cognitive impairment (MCI) and have acceptable quality neuroimaging. The ADNI cohort is significantly older (mean of 72.3 years versus 63.9 in PPMI).

We also train a regression model on the matched subjects to adjust imaging variables for systematic differences dut to study populations (ADNI vs PPMI), MRI manufacturers (‘GE’, ‘Philips’, ‘Siemens’) and magnetic field strength. Control subjects aged between 50 and 70 are designated as training samples. The regression map is learned and applied to each IDP throughout the full cohort using the `subtype`r function `adjustByCovariates`. The purpose of this process is to mitigate the influence of different imaging protocols, ensuring that subsequent analyses are less confounded by these factors. This approach has been used in practical studies of ADNI MRI data (Risacher et al. 2017). These merged and adjusted IDP data records are included in the file `ppmi_idps_trim_v1.4.0_SRF.csv`. SiMLR derived variables are denoted `t1PC*` (left-right averaged T1w derived feature sets), `t1aPC*` (asymmetry-related T1w derived feature sets), `dtPC*` (left-right averaged DTI derived feature sets), `dtaPC*` (asymmetry-related DTI derived feature sets) and `rsfPC*` (resting connectivity) where the `*` varies from 1 to 100. These SiMLR derived variables limit the multiple testing considerations to 100 variables because these are typically grouped together. That is, a given PC set (referred to as *simIDP ^k^*) is included in a single model (e.g. if *k* = 1, then *age ≈ t*1*PC*1+*t*1*aPC*1+*dt*1*PC*1+*dtaPC*1+*rsf PC*1) where *simIDP*_1_ = *t*1*, simIDP*_2_ = *t*1*a*, etc. We use this approach in a technical validation section below.

### Technical Validation

Components of technical validity that are critical for quantitative methodology in neuroimaging include: (a) generally robust performance across modalities; (b) multi-site reproducibility; (c) disease-specific discrimination from controls in particular over time in the clinical trial setting; (d) sensitivity to or relationship with changes in clinically relevant symptoms at baseline and/or over time. We provide evidence that the current IDPs satisfy these properties in the following sections.

- We quantify reproducibility and reliability in each modality through analysis of three traveling subject cohorts (addressing (a) and (b) above). These cohorts collect imaging data at different sites from the same individuals. Reliability data based on such cohorts are highly relevant for multisite trials which are always impacted by site-specific variation. By aggregating data from the traveling subject cohorts, we offer precise, reproducible reliability estimates (via intra-class correlation) manifested across different scanner types and imaging modalities.
- We derive effect sizes from statistical models that test established hypotheses comparing biomarker classified PD subjects versus control subjects. These show expected effects of PD are detectable in these data. This addresses (c) above.
- We finalize the technical validity section with examples of how scientists may relate IDP measures of brain health to rate of symptom change in PD in a multiple modality (integrative) context. This addresses (d) above.

The scale of the current data supports control for a subset of important PD relevant covariates including disease duration, educational level, sex, age and levodopa dose equivalent daily dose (LEDD). These variables are included in reference models with additional details below.

### Robust performance

The technical validity of these methods is supported by previous work, including various open quantitative MRI analysis challenges (Menze et al. 2015; Murphy et al. 2011; Baheti et al. 2021) that span modalities and organ systems. The foundational methods also support applications to non-human data (Allan Johnson et al. 2019; Hopkins and Avants 2013). Furthermore, the consistency of the methodology naturally enables multivariate statistical inference and/or prediction (Stone et al. 2023) even within the multi-study context (Dadu et al. 2024).

### Multi-site reproducibility

Traveling subject studies involve scanning the same subjects on multiple MRI scanners at different locations. These studies help in assessing consistency and/or agreement of image quantification where the only variables are the machines themselves. This is crucial for understanding power in multi-site studies of natural history or intervention and for ensuring that the observed changes in brain structure or function are due to actual physiological changes rather than variations in the imaging process itself.

In this study, we employ traveling cohort data (Hawco et al. 2022; Tanaka et al. 2021; Tong et al. 2019) to assess the agreement of IDPs pooled across multiple sites for the purposes of statistical inference. These data will establish expectations of repeatability for T1w, DTI and rsfMRI as measured by ANTsPyMM processing and are described briefly here:

1. the SRPBS Traveling Subject MRI Dataset (Tanaka et al. 2021):

- 9 healthy subjects travel to 12 sites to be imaged;
- of the 12 sites, 9 have consistently available T1w and rsfMRI in 6 subjects.
2. traveling subject DTI cohort (Tong et al. 2019):

- 3 healthy subjects travel to 4 sites to be imaged;
- T1w and multi-shell DWI/DTI are available.
3. Hawco’s traveling subject MRI dataset (Hawco et al. 2022) is available here:

- 4 healthy male subjects travel to 6 sites to be imaged with T1w, rsfMRI and DTI.

Thus, we use these data to characterize the consistency and reliability of these tools when applied to data that has known systematic biases due to site and scanner differences. The results confirm that findings and conclusions drawn from ANTsPyMM are reliable and not overwhelmed by scanner-specific differences or inconsistencies. This knowledge is critical for a foundational framework such as ANTsX/ANTsPyMM upon which scientific studies, machine learning platforms and other methodological comparisons are based. These cohorts represent variability in both MRI manufacturer and MRI model (high variability) that would exceed standard (within-scanner, within-site) test-retest analysis. Results therefore provide a lower-bound on reliability; i.e. within-site (e.g. longitudinal) studies would be expected to have higher reliability in general.

We employ the intra-class correlation to assess results. ICC ranges may be interpreted as (Koo and Li 2016):

- below 0.50: poor
- between 0.50 and 0.75: moderate
- between 0.75 and 0.90: good
- above 0.90: excellent

We find that ANTsPyMM IDPs derived from the same subjects imaged at different sites with MRI from various manufacturers show overall good to high reliability with a few exceptions within resting state derivatives (fALFF specifically). This provides empirical evidence that multiple modality MRI may be used to derive quantitative phenotypes on which predictive models may be based. Statistical control for site effects should still be applied at the population level using, for example, random effects. The data and code for reproducing these results is available in this location. Figure 7 shows the key summary output for this ICC comparison.

**Figure 7:**
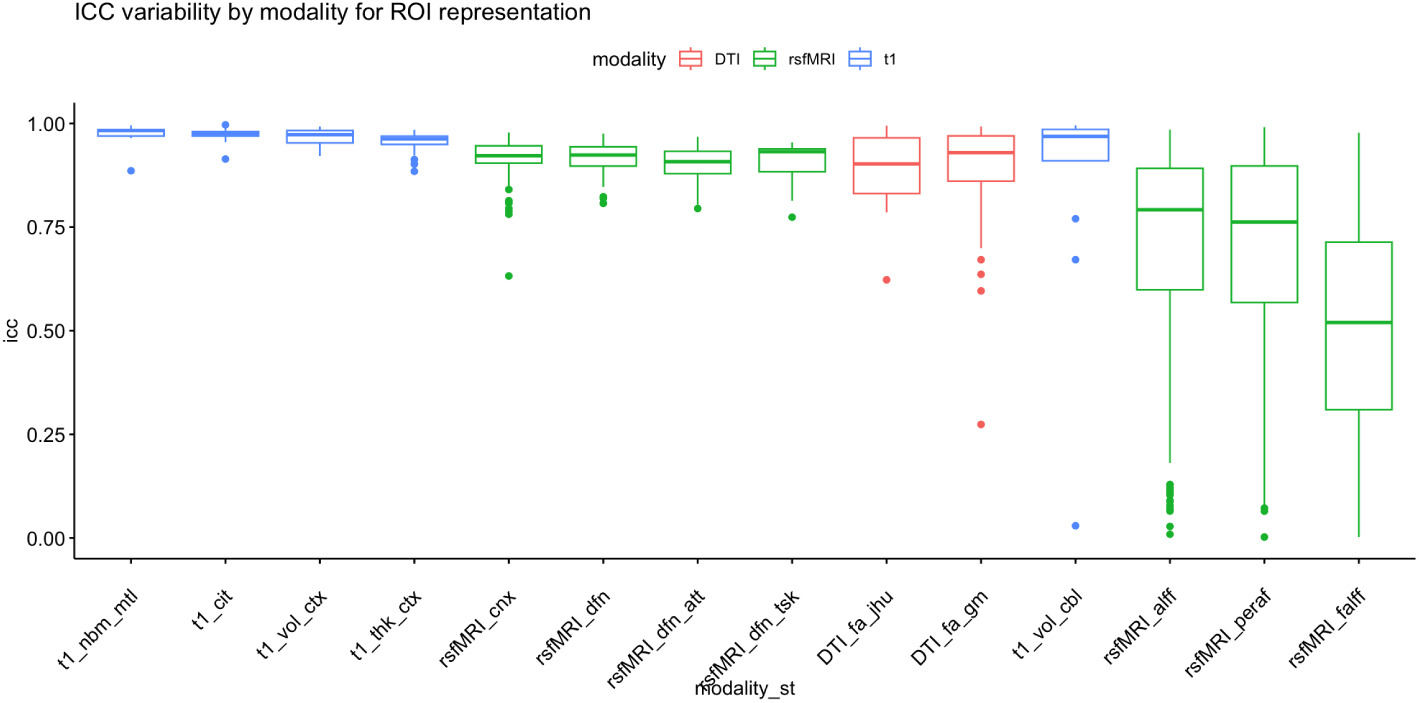
Summary reproducibility results from aggregated traveling subject data. T1 IDPs represent high reproducibility in all categories (cerebellum, CIT168, cortical volume, cortical thickness, basal forebrain and medial temporal lobe). DTI IDPs are also highly reproducible with FA in the cortical gray matter (gm) nearly equaling that of major white matter regions in the JHU atlas. Resting state connectivity shows good to excellent reproducibility; PerAF, fALFF and ALFF are relatively less reproducible – on average – though variability across regions is also high.

### Diagnostic effects in pre-defined structural, white matter and resting functional measurements

**Table 2.**
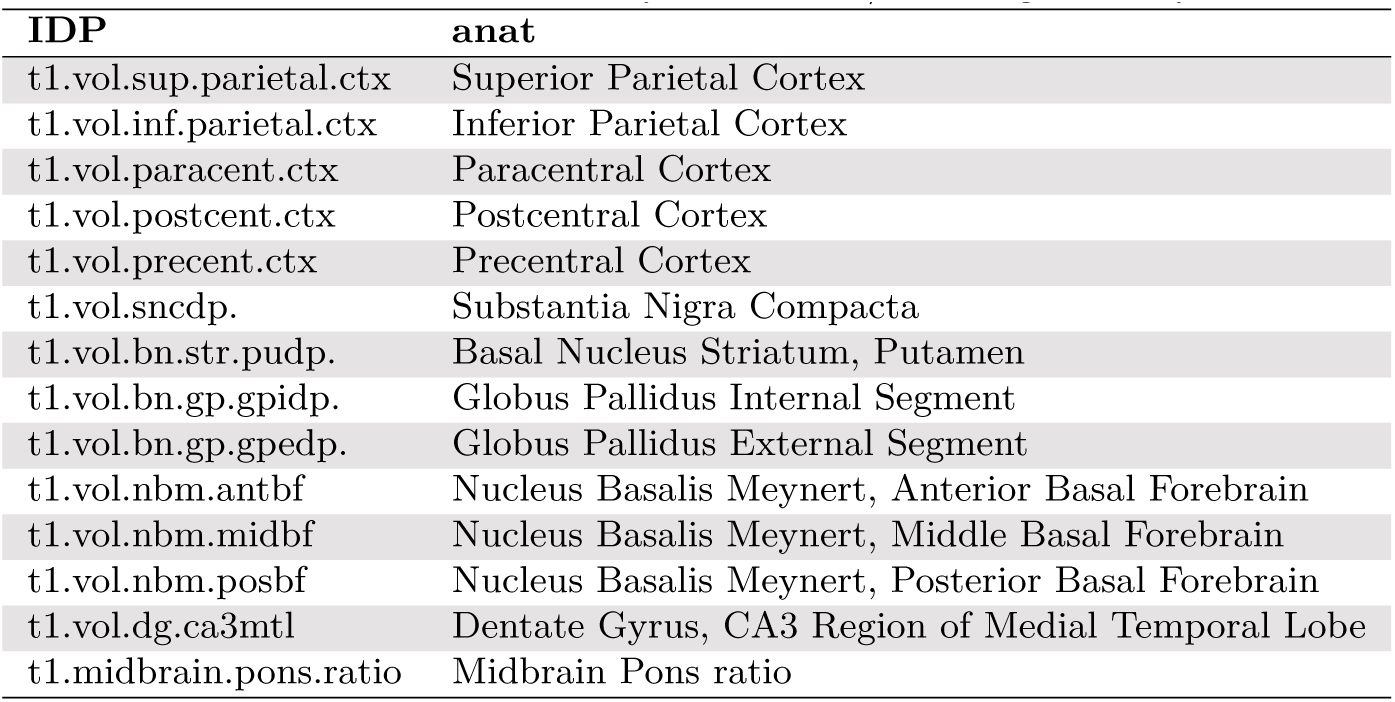
PD IDPs in ANTsPyMM: T1w L/R average and asym.

| IDP | anat |
| --- | --- |
| t1.vol.sup.parietal.ctx | Superior Parietal Cortex |
| t1.vol.inf.parietal.ctx | Inferior Parietal Cortex |
| t1.vol.paracent.ctx | Paracentral Cortex |
| t1.vol.postcent.ctx | Postcentral Cortex |
| t1.vol.precent.ctx | Precentral Cortex |
| t1.vol.sncdp. | Substantia Nigra Compacta |
| t1.vol.bn.str.pudp. | Basal Nucleus Striatum, Putamen |
| t1.vol.bn.gp.gpidp. | Globus Pallidus Internal Segment |
| t1.vol.bn.gp.gpedp. | Globus Pallidus External Segment |
| t1.vol.nbm.antbf | Nucleus Basalis Meynert, Anterior Basal Forebrain |
| t1.vol.nbm.midbf | Nucleus Basalis Meynert, Middle Basal Forebrain |
| t1.vol.nbm.posbf | Nucleus Basalis Meynert, Posterior Basal Forebrain |
| t1.vol.dg.ca3mtl | Dentate Gyrus, CA3 Region of Medial Temporal Lobe |
| t1.midbrain.pons.ratio | Midbrain Pons ratio |

**Table 3.**
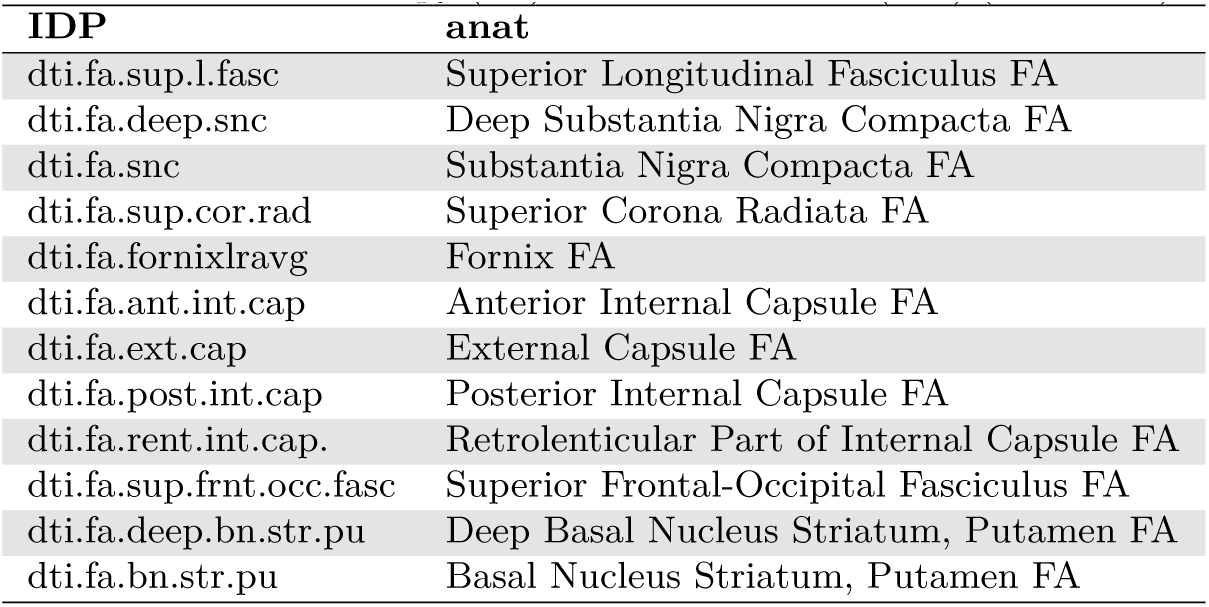
PD IDPs in ANTsPyMM: DTI L/R average and asym for both fractional anisotropy (FA) and mean diffusion (MD) (not shown).

**Table 4.** PD IDPs in ANTsPyMM: rsfMRI bilateral inter or intra-network connectivity (Yan, et. al. homotopic parcellation nomenclature).

| IDP | connectivity |
| --- | --- |
| rsf.p2.sommotb.2.temppar | Temporal Parietal Region |
| rsf.p2.sommotb.2.contc | Control Network Component C |
| rsf.p2.sommotb.2.contb | Control Network Component B |
| rsf.p2.sommotb.2.conta | Control Network Component A |
| rsf.p2.sommotb.2.sommotb | Somatomotor Area B |
| rsf.p2.sommotb.2.sommota | Somatomotor Area A |
| rsf.p2.sommotb.2.visperi | Peripheral Visual Area |
| rsf.p2.sommotb.2.viscent | Central Visual Area |
| rsf.p2.sommotb.2.striatum | Striatum |
| rsf.p2.sommotb.2.dopamine | Dopaminergic system |
| rsf.p2.sommotb.2.basalganglia | Basal Ganglia |
| rsf.p2.sommotb.2.midbrain | Midbrain |
| rsf.p2.sommota.2.temppar | Temporal Parietal Region |
| rsf.p2.sommota.2.contc | Control Network Component C |
| rsf.p2.sommota.2.contb | Control Network Component B |
| rsf.p2.sommota.2.conta | Control Network Component A |
| rsf.p2.sommota.2.sommotb | Somatomotor Area B |
| rsf.p2.sommota.2.sommota | Somatomotor Area A |
| rsf.p2.sommota.2.visperi | Peripheral Visual Area |
| rsf.p2.sommota.2.viscent | Central Visual Area |
| rsf.p2.sommota.2.striatum | Striatum |
| rsf.p2.sommota.2.dopamine | Dopaminergic system |
| rsf.p2.sommota.2.basalganglia | Basal Ganglia |
| rsf.p2.sommota.2.midbrain | Midbrain |

### Sensitivity to differences from controls

Symptoms of Parkinson’s disease (PD) exhibit heterogeneity both across individuals and within a single patient over time. MRI is suited to objective *in vivo* characterization of the neural basis of these changes longitudinally with a variety of structural and functional measurements. Here, we assess longitudinal and cross-sectional effect sizes in T1w, DTI and rsfMRI IDPs that are pre-defined for PD relevance. These regions are listed in Table 2 (T1w), Table 3 (DTI) and Table 4 (rsfMRI) and span motor, associative and limbic systems that may be impacted in PD and associated disorders (Ryman and Poston 2020). These include motor and parietal cortex (Filippi et al. 2020; Sokołowski et al. 2024), midbrain and striatal regions and basal forebrain (Batzu et al. 2023) from T1w. Due to known concomitant, AD-related pathology in some PD subjects, we also include a medial temporal lobe IDP (Das, Hwang, and Poston 2019). Relatedly, we select mean diffusion and FA derived from DTI in the striatum and substantia nigra (Hu et al. 2023) as well as major white matter tracts (Gattellaro et al. 2009; Pietracupa et al. 2018), the fornix and external and internal capsule; a recent large-scale study demonstrated sensitivity of these measures to PD (Owens-Walton et al. 2024). Interestingly, DTI metrics in early PD – within both the current study and the recent worldwide study (Owens-Walton et al. 2024) – appear to trend in directions that are opposite to that of other neurodegenerative diseases which provides an interesting opportunity for future work and more nuanced stage-based statistical modeling. In rsfMRI, we focus on connectivity between sensorimotor regions and other networks, in particular visual and cognitive control (Caspers et al. 2021; Wang et al. 2021; Tahmasian et al. 2015). In total, we test 99 different measurements which include left-right averaged as well as asymmetry metrics:

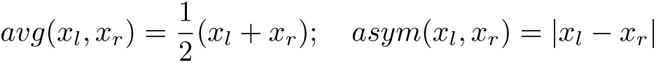

derived from those regions which are bilateral. This strategy is a generalizable way of testing laterality effects across all groups, including those without an established dominant side of disease (controls and pre-symptomatic PD subjects).

We provide exemplar linear mixed-effects models (LMMs) that seek to elucidate the complex relationships between neuroimaging biomarkers and the progression of Parkinson’s Disease (PD). The analytical framework was constructed using the R programming environment, leveraging the `lme4` package (Kuznetsova, Brockhoff, and Christensen 2017; Bates et al. 2014). This methodology allows for the exploration of hierarchical data structures commonly encountered in longitudinal neuroimaging studies, where multiple observations per subject are standard. These models were designed to investigate differences between biomarker-confirmed PPMI PD groups and controls. We accentuate that these models are for demonstration only and do not constitute fully-vetted “official” PPMI results. While these models may not account for all relevant covariates, they do provide evidence of validity by confirming the sensitivity of these IDPs to diagnostic group differences. These models are of the form:

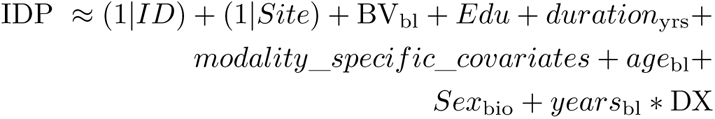

where IDP is the imaging outcome, *ID* is a random effect for subject ID, *Site* is a random effect for data collection site, BV_bl_ is baseline brain volume, *Edu* is educational attainment and *modality*_*specif ic*_*covariates* covaries for modalityspecific variability due to site and related effects. Age at baseline and biological sex are additional covariates. The primary predictors of interest are *years*_bl_ *∗* DX i.e. the interaction between time from baseline of the IDP measurement (in years) and the diagnostic group which includes SAA positive and negative PD subjects in addition to controls and MCI. The cross-sectional/longitudinal sample sizes and results for each group within the *DX* variable are denoted in Table 5 (cross-sectional) and 6 (longitudinal) where a postfix of “.x” indicates a cross-sectional estimate and “.y” a longitudinal one. The columns anv.x and anv.y reflect omnibus model *p*-values; `d` indicates effect size for the corresponding group either cross-sectionally or longitudinally. The `sig` column indicates whether the corrected *p*-value survives family-wise error (fwe) or false discovery rate (fdr) correction. In the longitudinal cohort, subjects are required to have two or more visits.

**Table 5.** Time independent group effects in PPMI/ADNI. Top-k per modality.

| voi | n.x (CN,Sp-,Sp+) | anv.x | d.x.Sp- | d.x.Sp+ | sig.x | sig.y |
| --- | --- | --- | --- | --- | --- | --- |
| t1.vol.bn.str.pudp. | 747/44/574 | p = 6.4813e-06 | -0.27 | -0.08 | fwe.x | fwe.y |
| t1.midbrain.pons.ratio | 747/44/574 | p = 6.6242e-08 | -0.23 | -0.26 | fwe.x | fwe.y |
| t1.vol.sncdp. | 747/44/574 | p = 2.2643e-16 | -0.41 | -0.30 | fwe.x | fwe.y |
| t1.vol.bn.gp.gpidp. | 747/44/574 | p = 4.5784e-18 | -0.47 | 0.05 | fwe.x | fdr.y |
| t1.vol.bn.gp.gpe.asymdp. | 747/44/574 | p = 3.6180e-07 | 0.29 | 0.13 | fwe.x | fdr.y |
| t1.vol.bn.gp.gpi.asymdp. | 747/44/574 | p = 2.0369e-06 | 0.27 | -0.01 | fwe.x | fdr.y |
| t1.vol.bn.str.pu.asymdp. | 747/44/574 | p = 7.2291e-22 | 0.54 | 0.04 | fwe.x | n.s. |
| t1.vol.bn.gp.gpedp. | 747/44/574 | p = 4.1291e-17 | -0.33 | 0.27 | fwe.x | n.s. |
| t1.vol.snc.asymdp. | 747/44/574 | p = 0.0004 | 0.21 | 0.01 | fwe.x | n.s. |
| t1.vol.nbm.midbf | 747/44/574 | p = 0.0002 | -0.23 | -0.06 | fwe.x | n.s. |
| t1.vol.nbm.antbf | 747/44/574 | p = 0.0022 | -0.18 | 0.01 | fdr.x | n.s. |
| dti.mean.md.snc | 327/29/365 | p = 0.0002 | 0.07 | 0.31 | fwe.x | fwe.y |
| dti.mean.md.deep.snc | 327/29/365 | p = 0.0005 | 0.06 | 0.29 | fwe.x | fwe.y |
| dti.fa.bn.str.pu | 327/29/365 | p = 0.0029 | 0.02 | -0.24 | fdr.x | fdr.y |
| rsf.p2.sommotb.2.striatum | 327/26/277 | p = 0.0035 | 0.20 | 0.23 | fdr.x | n.s. |

regression (`lm` in `R`) with the same structure as described above but without random effects. Linear mixed-effects models (LMMs) were constructed using the `lmer` function from the `lme4` package and fitted to the data. This process included the standardization of variables within the equation, a critical step given the varying scales and distributions of neuroimaging metrics. Comparative model analysis was conducted to ascertain the significance of various predictors, employing the anova function to contrast models with and without *DX* and the interaction between *DX* and *years*_bl_. This `anova` assesses the “omnibus” model improvement due to the joint addition of both *DX* and the interaction between *DX* and *years*_bl_. As we are only reporting high level results here, we do not investigate *p*-values within individual diagnostic groups. These results are shown in Table 5 (for the *DX* term) and Table 6 (for the longitudinal term). Effect sizes (d.x and d.y) for each term are estimated from the *t−*value and degrees of freedom for each cross-sectional and longitudinal model. Very small effect sizes are those with absolute value less than 0.2. The effects of *DX* on these IDPs are visualized through predictor effect plots (Larsen and McCleary 1972; Fox and Weisberg 2018) of diagnosis by time generated for each diagnostic category as in Figure 8. These IDPs are significant under family wise error (fwe) multiple comparisons correction at *p*-value *≤* 0.05 (after correction based on the above-mentioned `anova`). These plots and other regression plots are displayed with the `R` packages `jtools` and `interactions`.

**Figure 8:**
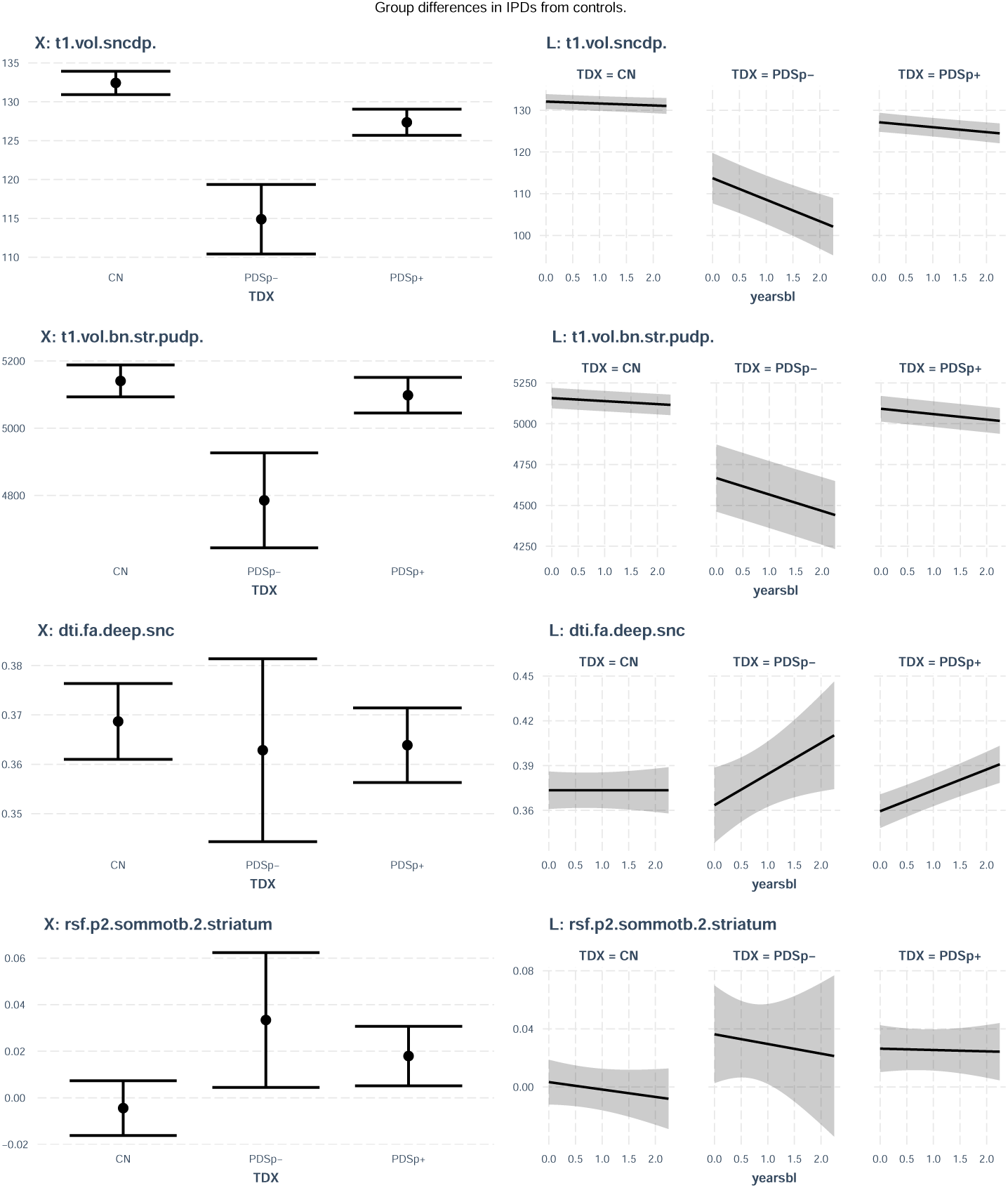
Partial regression plots for example significant IDPs illustrate the trends of differences from controls. The bar plots at left show 95 percent confidence intervals along with estimated means for each group. The line plots at right show the estimated change over time (0 to 4 years) along with 95 percent confidence intervals. Table 5 and 6 detail the associated significance levels and effect sizes.

**Table 6.**
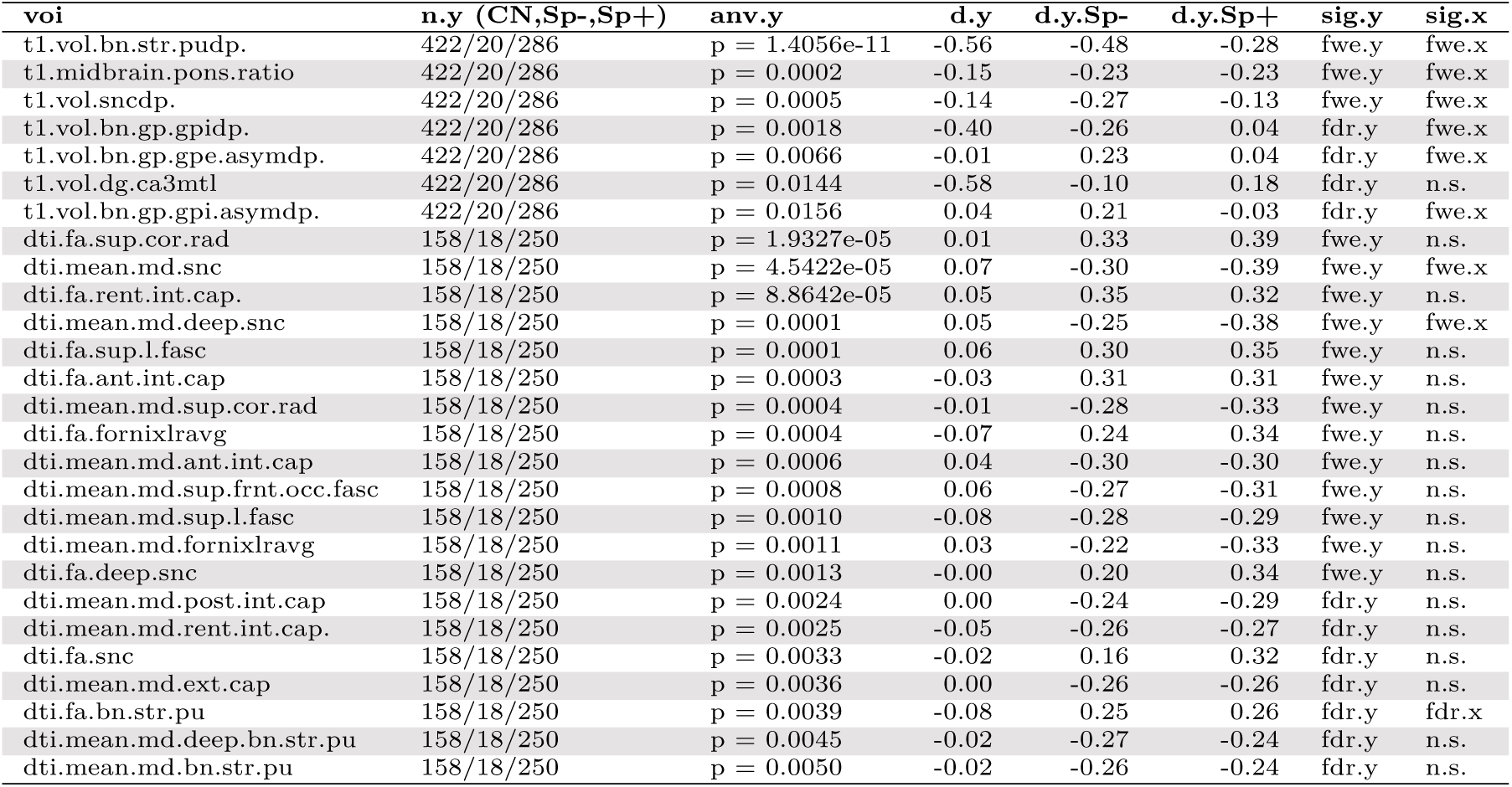
Longitudinal group effects in PPMI/ADNI. Top-k per modality.

### Baseline IDP to longitudinal MDS-UPDRS effects in structural, white matter and resting functional measurements

We use LMMs to estimate the relationship of IDP values to clinical observations as evaluated by MDS-UPDRS 1, 2, 3 (off), total (off) and related scores. While these clinical measurements have well-documented limitations in terms of reliability and interpretability (on behalf of the Parkinson’s Progression Markers Initiative et al. 2023), they are consistently available in PPMI. These exemplar assessments differ from the prior section in that they focus only on PPMI subjects as these measurements are absent in ADNI. These models are of the form:

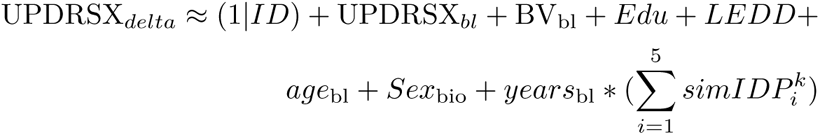

The outcome is UPDRSX*_delta_* indicating change in a given MDS-UPDRS score. The majority of these variables are as defined previously. However, we introduce covariates for treatment effects (Levodopa Equivalent Daily Dose, LEDD) as well as baseline values of the given score. This latter variable approximates a control for subject and domain-specific disease severity. As such, these models are relatively conservative in terms of their attribution of variance to IDP values. The predictor of interest, here, is *years*_bl_ *∗* 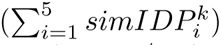 which estimates change in the given MDS-UPDRS score (or derived score / subscore) in relation to brain structure (including asymmetry) and function as measured by three modalities. The *p*-value associated with each of the *k* in 1 *· · ·* 100 models is determined by the amount of additional variance that is explained by the SiMLR IDPs (R anova function). We determine significance from the omnibus *p*-value returned by anova. The effect sizes are derived from the single most predictive SiMLR IDP in each model; additional influence by secondary IDPs would augment estimates. These models are assessed in the range of baseline to 2.25 years change. As in the prior section, cross-sectional results are derived with standard regression (lm); the outcome in this case, however, is the raw score, not its change.

The effects are visualized, in Figure 9, through effect plots for select IDPs of interest. Effect sizes for each significant (fdr or fwe) pair of outcomes and IDPs in Table 7 where S.m1.1 indicates the top IDP feature contributing to the model. If S.m2.1 is present, this means that a second predictor also contributes significantly to the association (uncorrected *p ≤* 0.05 based on the *t*-value for a given 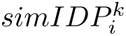 where *i* indexes the individual type of measurement and *k* indicates the *k*^th^ component). The clinical measurement and its most highly weighted IDP is also listed. In several cases, two or more distinct modalities contribute effectively. Moreover, although SiMLR’s multivariate feature learning was performed on a separate cohort, several reasonable associations are brought out by this analysis: involvement of hypothalamus FA (`dti.fa.die.hth`) with REM sleep disturbance (rem), connectivity between dopaminergic regions and cognitive control regions (`rsf.p2.conta.2.dopamine`) and MDS-UPDRS-III scores and thickness of substantia nigra pars compacta (`t1.thk.sncdp`) with several measures including postural instability and gait disturbance (PIGD). Recall that we are only reporting, in Table 7, the top features; each 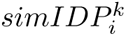 involves many regions. Region names are described in detail online in the data dictionary and associated documentation.

**Figure 9:**
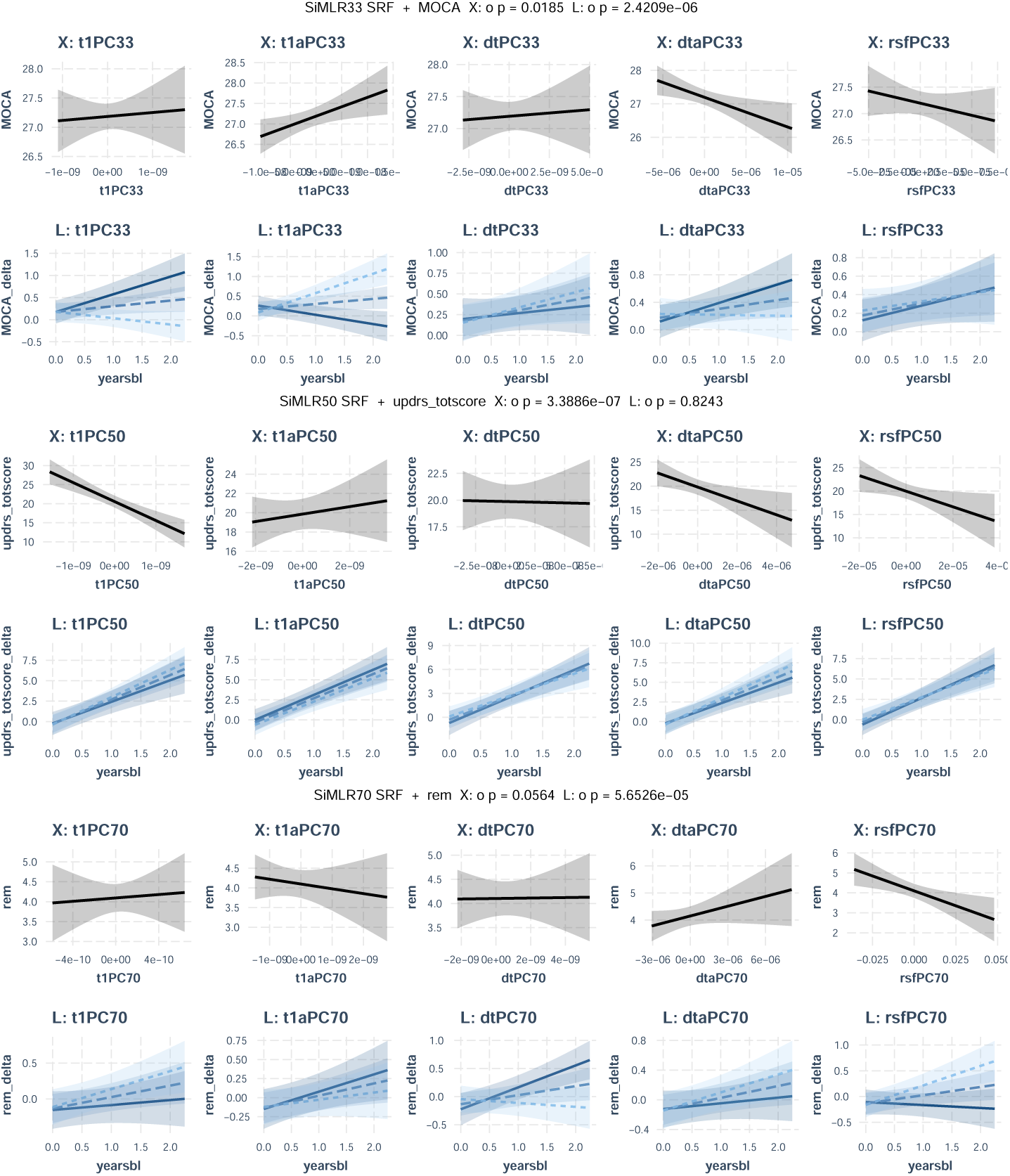
Select predictor effect plots for SiMLR mapping between M3RI and PD symptomology. Plots headed with “X” relate to cross-sectional effects while “L” is longitudinal. Significance for the class of effects is also noted in the main title of each figure pair (“p = . . . .”). Shaded regions in all panels show 95 percent confidence intervals. In the “L” plots, the predictor effect plots visualizes interaction between time from baseline and the given SiMLR IDP. The darker lines indicate the relationship of higher values in the given imaging score with the change in the outcome. Lighter dashed lines indicate the relationship of lower values in the given imaging score with the change in the outcome. For example, higher values in t1PC33 are associated with MOCA preservation over time (or increased learning) as is decreased asymmetry. The SiMLR50 plots (middle panels) show robust cross-sectional association with total score but no evidence of longitudinal association. The SiMLR70 plots (bottom pair) show association of both DTI and resting s2t5ate IDPs with REM sleep disturbance changes over time.

**Table 7.**
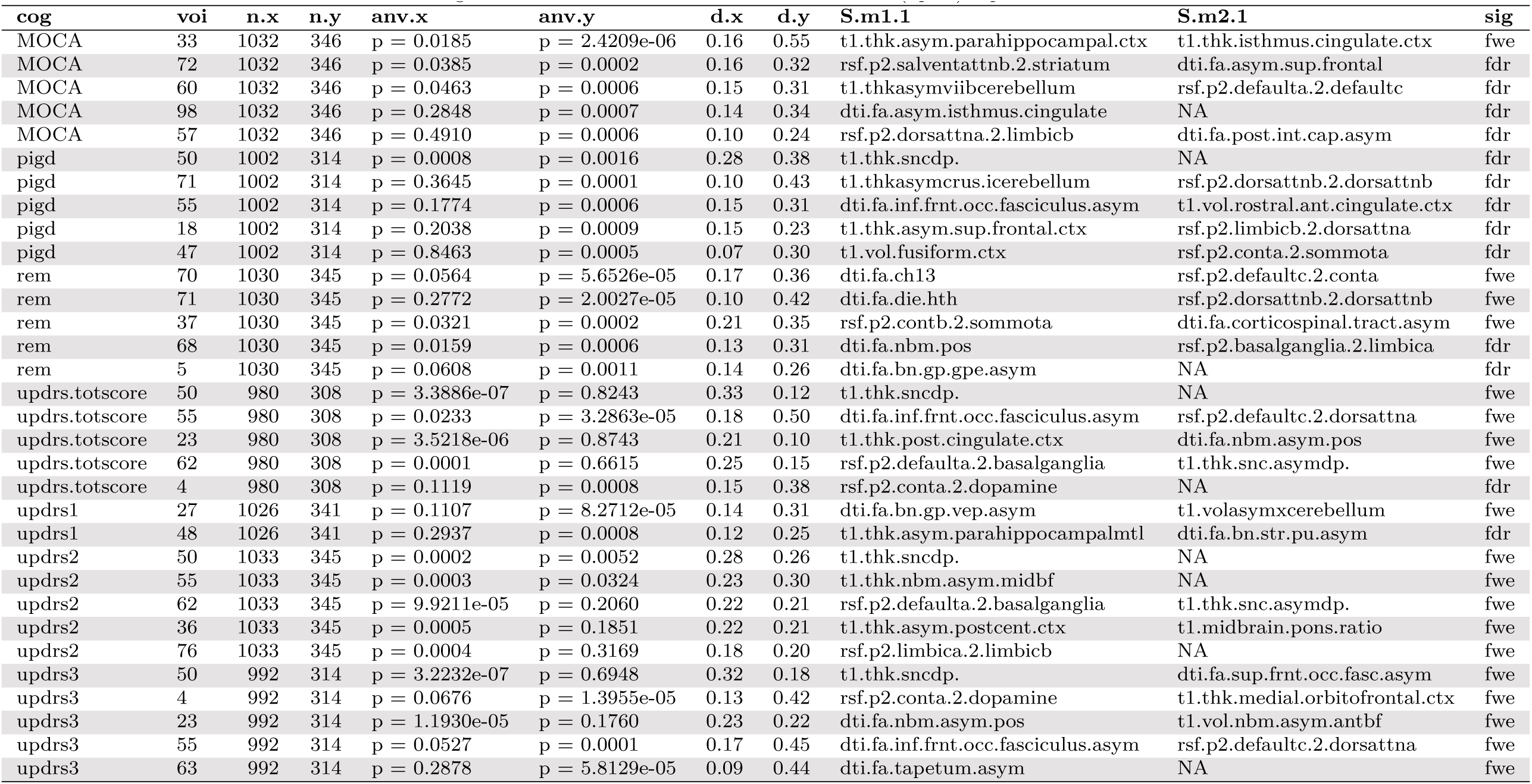
Significant SiMLR IDP to clinical measurements: (up to) top five for each score.

This SiMLR study demonstrates that brain state – as measured jointly by these modalities – may contribute to acceleration/deceleration of changes in MDS-UPDRS and related scores. However, we accentuate that these models are relatively simple and linear; as such, they yield only rough suggestions that additional modeling effort may be warranted to understand the differential value of these IDPs across the spectrum of PD symptomology. Furthermore, this analysis grouped all PPMI subjects together (excluding controls); as such, it lacks the specificity that may be needed to parse subgroup relationships or those that only occur within specific stages of PD. The subjects comprising these results span a variety of PD-related subgroups: PDGBA: 8 / PDLRRK2: 14 / PDSNCA: 1 / PDSporadic: 367 / ProdromalGBA: 8 / ProdromalLRRK2: 24 / ProdromalSporadic: 533 (median across results for the baseline cohort; the longitudinal cohort subjects are fewer as they are required to have two or more visits: PDGBA: 6 / PDLRRK2: 7 / PDSporadic: 172 / ProdromalGBA: 7 / ProdromalLRRK2: 16 / ProdromalSporadic: 133). Despite these limitations, Table 7 demonstrates that all three modalities studied here may jointly influence clinical presentation and/or symptoms. Furthermore, several of these models indicate that multiple IDPs are changing in concert with symptom progression.

In summary, these data and reference results demonstrate the potential of multimodal neuroimaging and integrative statistical approaches in PD and neurodegenerative disease research more generally. This tabulated multi-modality MR IDP dataset for PPMI – derived from deeply validated open source methods – represents a valuable opportunity to help standardize as well as advance PD M3RI research. It simplifies – to the extent that is currently possible – the analysis of complex imaging data and potentially accelerates the discovery of novel insights into PD progression and effects. The timing of this data release is critical given the newly available SAA biomarker. Additionally, this methodology holds promise for broader applications, potentially benefiting research into other neurological conditions.

### Usage Notes

An example of processing used here is shown in the github respository https://gi thub.com/stnava/ANTPD_antspymm where we combine easily accessible multiview neuroimaging with our open source methods for demonstration purposes. All images referred to in this research were processed in a style identical to this example.

## Code availability

Core image processing was done with `python 3.9` while document creation was achieved with `R` version `4.3.0` – “Already Tomorrow”. ANTsPyMM is installable via `pypi` and available at github. The version used for this work is `1.4.0` along with tensorflow `2.11.0`, antspyx `0.5.0`, antspynet `0.2.8` and antspyt1w `0.9.4`. Subtyper `v1.0.0` and ANTsR `0.6.2` contain many utilities used in the creation of this compilable document which was built with Rmarkdown (Xie, Dervieux, and Riederer 2020).

## Data Availability

All data produced in the present study are available upon reasonable request to the authors

https://ida.loni.usc.edu/login.jsp?project=PPMI

## Acknowledgements

Our sincere appreciation to the Michael J. Fox Foundation (MJFF) for supporting this work through MJFF-021144. Technical development for aspects of the software used in this work are supported by the Office of Naval Reseach ONR Award Number: N00014-23-1-2317.

## Author contributions statement

B.A. conceptualized and designed the study. L.F., O.H., A.R., A.S., X.W. conducted the experiments and collected the data. B.A. and L.B. performed data analysis and interpretation. N.J.T., J.R.S. and P.A.C. contributed to the development of the software framework and methodology. A.S. and A.R. were responsible for the software and computational tools used in the study. L.C., B.M., R.G, K.P. and K.M. supervised the research and provided critical feedback on the manuscript. B.A. wrote the first draft of the manuscript. All authors reviewed and approved the final version of the manuscript.

