## supplemental information for "Magnetic Resonance Imaging Data Phenotypes for the Parkinson’s Progression Markers Initiative"

---

*Data used in preparation of this article were obtained from the Alzheimer's Disease Neuroimaging Initiative (ADNI) database (adni.loni.usc.edu). As such, the investigators within the ADNI contributed to the design and implementation of ADNI and/or provided data but did not participate in analysis or writing of this report. A complete listing of ADNI investigators can be found at: http://adni.loni.usc.edu/wp-content/uploads/how_to_apply/ADNI_Acknowledgement_List.pdf

LaTeX{ }

### Overview of the supplement

We first provide additional information on the currently processed ADNI and PPMI cohort including the genetic and prodromal subgroups along with asyn SAA status for each. Second, we share didactic material explaining in greater detail how to interpret the plots in Figure 9 of the main text.

#### The full PPMI cohort

The main text focused on the largest groups available in this data release: controls and sporadic PD. However, other PD groups are of growing interest for more focused research. In particular, both GBA and LRRK2 PD appear to take a different disease trajectory than is typical of sporadic PD [1] Moreover, the prodromal stage is of growing interest due to the combination of an increased ability to identify these subjects and the potential for early intervention. We believe the validity arguments made in the primary text extends to the IPDs that are associated with these other groups, summarized in Table S1.

LaTeX{ }

[1]Typical should probably not be used to describe PD progression due to the immense variability that is observed across individual patients.

Table S1. Baseline PPMI IDP cohort. Pro=prodromal; MSA = multi-system atrophy; OtherGen = other genetic; RBD = REM Sleep Behavior Disorder.

| | **CN (N=748)** | **Gen.Pro (N=553)** | **Sporadic.Pro (N=811)** | **OtherGen.PD (N=10)** | **LRRK2.PD (N=185)** | **GBA.PD (N=119)** | **Sporadic.PD (N=851)** | p |
|---|---|---|---|---|---|---|---|---|
| age | 69.8 ± 9.8 | 62.3 ± 7.4 | 68.1 ± 5.6 | 49.9 ± 14.0 | 64.9 ± 8.5 | 62.2 ± 9.6 | 63.6 ± 9.2 | < 0.001 |
| Sex | | | | | | | | < 0.001 |
| Female | 356 (47.6%) | 331 (59.9%) | 400 (49.3%) | 2 (20.0%) | 98 (53.0%) | 54 (45.4%) | 308 (36.2%) | |
| Male | 392 (52.4%) | 222 (40.1%) | 411 (50.7%) | 8 (80.0%) | 87 (47.0%) | 65 (54.6%) | 543 (63.8%) | |
| race | | | | | | | | < 0.001 |
| Asian | 4 ( 0.5%) | 0 ( 0.0%) | 3 ( 0.4%) | 0 ( 0.0%) | 1 ( 0.5%) | 0 ( 0.0%) | 14 ( 1.6%) | |
| Black | 10 ( 1.3%) | 0 ( 0.0%) | 6 ( 0.7%) | 0 ( 0.0%) | 0 ( 0.0%) | 2 ( 1.7%) | 11 ( 1.3%) | |
| not.spec. | 507 (67.8%) | 6 ( 1.1%) | 8 ( 1.0%) | 0 ( 0.0%) | 0 ( 0.0%) | 0 ( 0.0%) | 6 ( 0.7%) | |
| Other | 8 ( 1.1%) | 12 ( 2.2%) | 27 ( 3.3%) | 0 ( 0.0%) | 14 ( 7.6%) | 2 ( 1.7%) | 24 ( 2.8%) | |
| White | 219 (29.3%) | 535 (96.7%) | 767 (94.6%) | 10 (100.0%) | 170 (91.9%) | 115 (96.6%) | 796 (93.5%) | |
| duration.yrs | | 0.8 ± 1.2 | | 3.6 ± 3.7 | 3.0 ± 2.1 | 2.7 ± 1.9 | 0.7 ± 0.6 | < 0.001 |
| updrs.totscore | 4.8 ± 4.4 | 9.5 ± 8.8 | 13.1 ± 10.4 | 33.1 ± 16.7 | 37.4 ± 17.7 | 45.5 ± 14.9 | 35.0 ± 15.1 | < 0.001 |
| CSFSAA | | | | | | | | < 0.001 |
| Negative | 474 (86.2%) | 484 (92.4%) | 127 (35.8%) | 4 (50.0%) | 61 (35.7%) | 6 ( 6.7%) | 49 ( 7.4%) | |
| Positive | 76 (13.8%) | 39 ( 7.4%) | 226 (63.7%) | 4 (50.0%) | 109 (63.7%) | 82 (91.1%) | 608 (92.3%) | |
| PosMSA | 0 ( 0.0%) | 1 ( 0.2%) | 2 ( 0.6%) | 0 ( 0.0%) | 1 ( 0.6%) | 2 ( 2.2%) | 2 ( 0.3%) | |
| LEDD | 0.0 ± 0.0 | 4.1 ± 45.8 | 0.4 ± 10.5 | 573.2 ± 450.4 | 465.0 ± 425.7 | 542.3 ± 551.3 | 9.9 ± 62.6 | < 0.001 |
| subgroup | | | | | | | | < 0.001 |
| GBA | 0 ( 0.0%) | 281 (50.8%) | 0 ( 0.0%) | 0 ( 0.0%) | 0 ( 0.0%) | 119 (100.0%) | 0 ( 0.0%) | |
| GBA + RBD | 0 ( 0.0%) | 1 ( 0.2%) | 0 ( 0.0%) | 0 ( 0.0%) | 0 ( 0.0%) | 0 ( 0.0%) | 0 ( 0.0%) | |
| Healthy Control | 242 (100.0%) | 0 ( 0.0%) | 0 ( 0.0%) | 0 ( 0.0%) | 0 ( 0.0%) | 0 ( 0.0%) | 0 ( 0.0%) | |
| Hyposmia | 0 ( 0.0%) | 0 ( 0.0%) | 567 (69.9%) | 0 ( 0.0%) | 0 ( 0.0%) | 0 ( 0.0%) | 0 ( 0.0%) | |
| LRRK2 | 0 ( 0.0%) | 245 (44.3%) | 0 ( 0.0%) | 0 ( 0.0%) | 178 (96.2%) | 0 ( 0.0%) | 0 ( 0.0%) | |
| LRRK2 + GBA | 0 ( 0.0%) | 24 ( 4.3%) | 0 ( 0.0%) | 0 ( 0.0%) | 7 ( 3.8%) | 0 ( 0.0%) | 0 ( 0.0%) | |
| PRKN | 0 ( 0.0%) | 0 ( 0.0%) | 0 ( 0.0%) | 5 (50.0%) | 0 ( 0.0%) | 0 ( 0.0%) | 0 ( 0.0%) | |
| RBD | 0 ( 0.0%) | 0 ( 0.0%) | 244 (30.1%) | 0 ( 0.0%) | 0 ( 0.0%) | 0 ( 0.0%) | 2 ( 0.2%) | |
| SNCA | 0 ( 0.0%) | 2 ( 0.4%) | 0 ( 0.0%) | 5 (50.0%) | 0 ( 0.0%) | 0 ( 0.0%) | 0 ( 0.0%) | |
| Sporadic PD | 0 ( 0.0%) | 0 ( 0.0%) | 0 ( 0.0%) | 0 ( 0.0%) | 0 ( 0.0%) | 0 ( 0.0%) | 849 (99.8%) | |
| imaging.protocol | | | | | | | | < 0.001 |
| ADNI | 506 (67.6%) | 0 ( 0.0%) | 0 ( 0.0%) | 0 ( 0.0%) | 0 ( 0.0%) | 0 ( 0.0%) | 0 ( 0.0%) | |
| PPMI1 | 169 (22.6%) | 538 (97.3%) | 60 ( 7.4%) | 3 (30.0%) | 172 (93.0%) | 110 (92.4%) | 388 (45.6%) | |
| PPMI2 | 73 ( 9.8%) | 15 ( 2.7%) | 751 (92.6%) | 7 (70.0%) | 13 ( 7.0%) | 9 ( 7.6%) | 463 (54.4%) | |

#### Understanding interaction effect plots

Interaction plots, such as those generated by the **`interactions`** package, help visualize the relationship between two or more predictor variables in a regression model, highlighting how the effect of one variable (the "moderator") changes depending on the value of another (the "focal" variable). In the manuscript, we employ the `interact_plot()` function to plot the interaction between **`yearsbl`** and a given IDP. These plots include confidence intervals to show the precision of the estimates.

The first plot faceted by the moderator (**`facet.modx = TRUE`**) presents multiple small plots, each corresponding to a different level of the moderator variable, allowing the reader to examine the interaction within each subgroup separately. This is helpful when the effect of the predictor might differ substantially across levels of the moderator. The second plot, with **`facet.modx = FALSE`**, integrates the interaction in a single plot, showing how the interaction unfolds across the entire range of the moderator. The absence of plotted points and partial residuals ensures that the reader focuses solely on the fitted lines and their confidence intervals, making it easier to interpret the overall trend without distraction from individual data points or noise.

The reader should interpret these interaction plots by comparing how the slope of **`yearsbl`** (the focal predictor) changes depending on different values of the moderator variable. A steeper or flatter slope in different facets (or sections of the integrated plot) suggests that the relationship between **`yearsbl`** and the outcome variable varies depending on the moderator, implying a significant interaction effect. The example shown below focuses on just one of the SiMLR IDPs and how it relates to the MDS-UPDRS total score. As in the main text, the "omnibus" effect is gained by using `anova` to comare a baseline model (no IDPs) to an extended model with IDPs. The *p*-value for this example is $p < .001$ which indicates that the additional IDP variables improve the model fit significantly. The coefficients for the base model are shown in Table S2. The coefficients for the extended model are shown in Table S3. Investigating the invididual *p*-values in the extendend model (Table S3) suggests that the asymmetry in the DTI-derived variables (dtaPC52) and the resting state connectivity variables (rsfPC52) are most responsible for this improvement. Figure 1 illustrates this visually for dtaPC52 while Figure 2 shows rsfPC52. The remainder of the effect plots in the main manuscript can be interpreted in the same or a similar manner.

#### The full SiMLR feature sets for these predictors

We can also show − here − with this additional space the full feature sets associated with these SiMLR IDPs. Figure 3 shows the asymmetry related features. Figure 4 shows the resting state features. A higher weight indicates a larger contribution to the final predictor value. Note that weights are unsigned and therefore represent an average of these variables. The maximum weight for each feature vector is 1 and we display only those features with individual

Table S2: Fixed effects from the baseline mixed effects model of total MDS-UPDRS score

| term | estimate | statistic | df | p.value |
|---|---|---|---|---|
| (Intercept) | -1.589 | -1.694 | 248 | 0.091 |
| updrs_totscore_BL | -1.471 | -3.144 | 193 | 0.002 |
| LEDD | -1.706 | -3.336 | 358 | 0.001 |
| commonEdu | 0.170 | 0.376 | 185 | 0.707 |
| age_BL | 0.546 | 1.165 | 184 | 0.246 |
| commonSexMale | 1.075 | 1.084 | 178 | 0.280 |
| yearsbl | 4.352 | 6.214 | 334 | 0.000 |

Table S3: Fixed effects from the extended (baseline + SiMLR IDP) model of total MDS-UPDRS score

| term | estimate | statistic | df | p.value |
|---|---|---|---|---|
| (Intercept) | -1.909 | -1.839 | 246 | 0.067 |
| updrs_totscore_BL | -1.519 | -3.292 | 187 | 0.001 |
| LEDD | -1.491 | -2.942 | 346 | 0.003 |
| commonEdu | 0.072 | 0.163 | 181 | 0.871 |
| age_BL | 1.271 | 2.035 | 179 | 0.043 |
| commonSexMale | 1.120 | 1.156 | 166 | 0.249 |
| yearsbl | 4.330 | 5.462 | 329 | 0.000 |
| t1PC52 | 11555.905 | 0.896 | 268 | 0.371 |
| t1aPC52 | 9838.937 | 0.587 | 316 | 0.557 |
| dtPC52 | 305.571 | 0.038 | 310 | 0.970 |
| dtaPC52 | -2297.966 | -0.399 | 334 | 0.690 |
| rsfPC52 | 358.339 | 0.302 | 316 | 0.763 |
| yearsbl:t1PC52 | 3401.415 | 0.344 | 299 | 0.731 |
| yearsbl:t1aPC52 | 9303.726 | 0.557 | 324 | 0.578 |
| yearsbl:dtPC52 | 9333.378 | 1.078 | 347 | 0.282 |
| yearsbl:dtaPC52 | 15601.782 | 2.546 | 343 | 0.011 |
| yearsbl:rsfPC52 | 4442.960 | 3.323 | 349 | 0.001 |

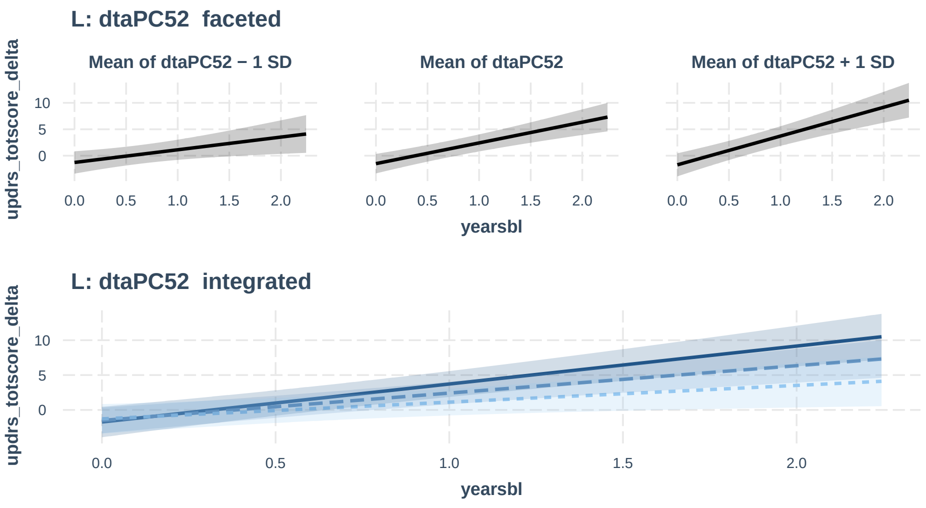


Figure 1: SiMLR mapping between M3RI and PD symptomology: DTA. The effect plot for the interaction of dtaPC52 with time shows its influence of UPDRS total score. The top plot shows the faceted version of the plot on the bottom. I.e. the same information is in both plots. We see that increased asymmetry in these dMRI-derived variables leads to increased change in the score over time.

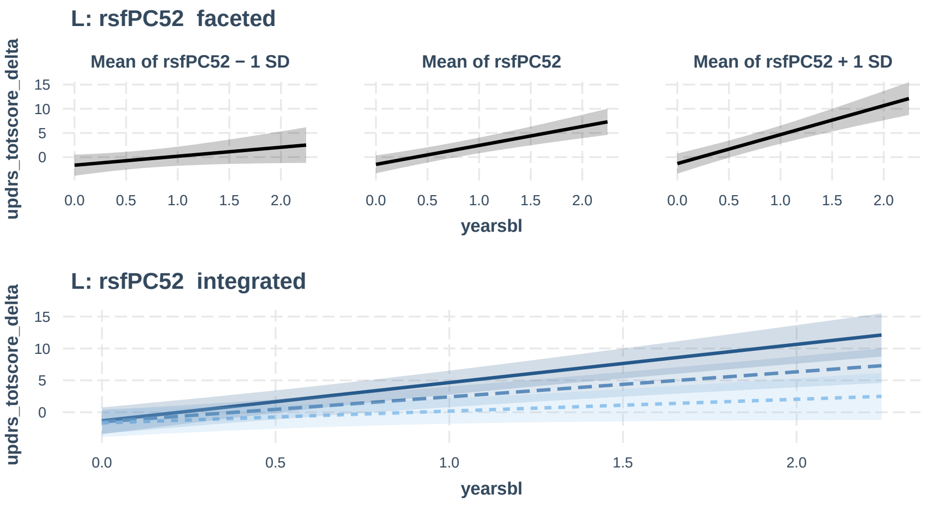


Figure 2: SiMLR mapping between M3RI and PD symptomology: rsfMRI. The effect plot for the interaction of dtaPC52 with time shows its influence of UPDRS total score. The top plot shows the faceted version of the plot on the bottom. I.e. the same information is in both plots. We see that increased connectivity in these rsfMRI-derived variables leads to increased change in the score over time.

weights greater than or equal to a given threshold. The explicit `ANTsR` code for these displays is shown.

```
n.comp=100
simfile=paste0("~/Downloads/ppmi_pym_data/pub_",n.comp,
  '_rezOR_only_ADNI')
presim=read_simlr_data_frames( path.expand(simfile),
      c("t1","t1a", "dt", "dta", "rsf" ))
ppmitrim = apply_simlr_matrices(  ppmitrim, presim, robust=FALSE,
  center = TRUE, scale = TRUE,
  absolute_value=rep(TRUE,length(presim)) )[[1]]
wmat=c("dta",52)
v0=interpret_simlr_vector2( presim[[wmat[1]]],
  paste0('PC',wmat[2]), n2show=50 )
v0=v0[v0>0.01]
todisp=data.frame( feature=names(v0), weight=v0 )
ggbarplot(todisp,'feature','weight') +
  ggplot2::theme( axis.text.x =
    ggplot2::element_text(angle = 80, hjust = 1))
```

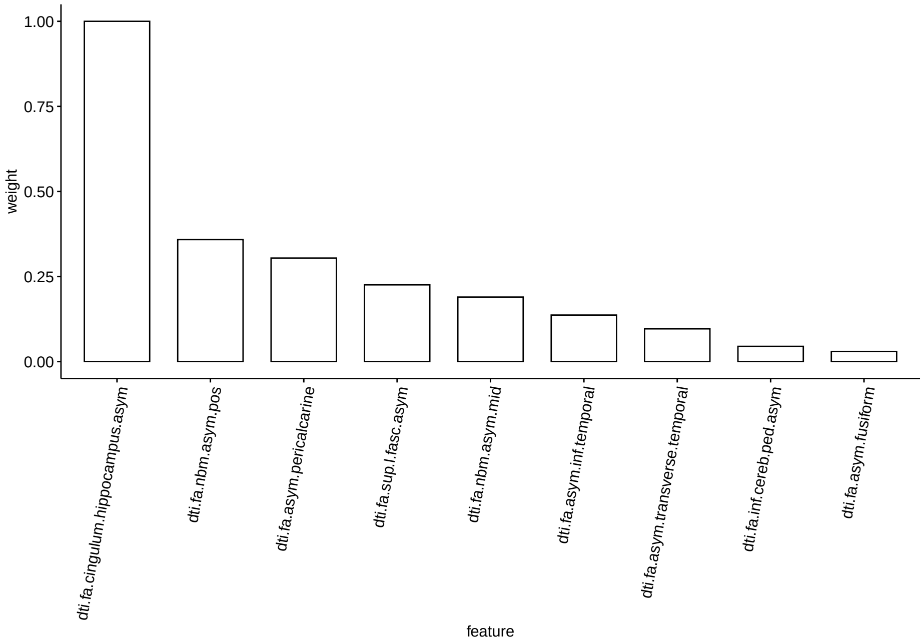


Figure 3: The most highly weighted features for dtaPC52.

```
wmat=c("rsf",52)
v1=interpret_simlr_vector2( presim[[wmat[1]]],
  paste0('PC',wmat[2]), n2show=80 )
```

```r
v1=v1[v1>0.1]
todisp=data.frame( feature=names(v1), weight=v1 )
ggbarplot(todisp,'feature','weight') +
  ggplot2::theme(axis.text.x =
    ggplot2::element_text(angle = 80, hjust = 1))
```

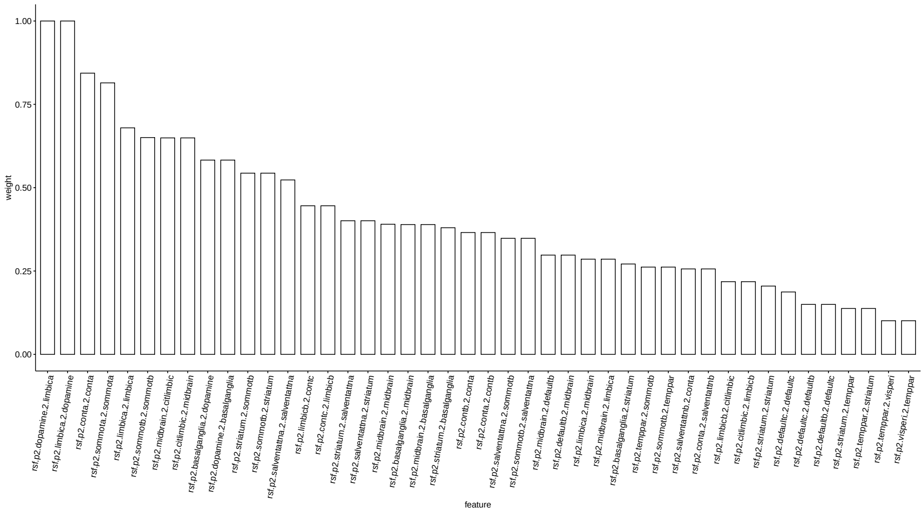


Figure 4: The most highly weighted features for rsfPC52.
